# A Sternum-Worn, Non-Invasive Vibrotactile Device to Alleviate Symptoms in Parkinson’s: a Multi-Site Double-Blind Randomised Controlled Trial

**DOI:** 10.1101/2025.07.19.25331818

**Authors:** Viktoria Azoidou, Essa Bhadra, Ellen Camboe, Kamalesh C. Dey, Alexandra Zirra, Corrine Quah, Caroline Budu, Thomas Boyle, David Gallagher, Jonathan P. Bestwick, Alastair J Noyce, Cristina Simonet

**Author notes:** These authors contributed equally to this work and share joint senior authorship as final authors (Alastair J Noyce, Cristina Simonet). Formatted in British English. Corresponding authors: Centre for Preventive Neurology, Wolfson Institute of Population Health, London, United Kingdom., &.

## Abstract

**Background:** Effective, measurable, and non-invasive treatments for motor symptoms, gait and balance difficulties, and associated non-motor burden in Parkinson’s disease (PD) remain limited. We aimed to assess the usability, safety/tolerability, and clinical efficacy of CUE1+, a wearable cueing and vibrotactile stimulation non-invasive device, in people with PD.

**Methods:** This 12-week, double-blind, randomised controlled trial was conducted at two UK sites in adults with idiopathic PD. Participants were randomly assigned (1:1) to receive either an active CUE1+ device or a sham device, worn on the sternum for 8 hours daily. Participants, investigators, and assessors were blinded to treatment allocation. The primary outcomes were usability and safety/tolerability of CUE1+. Secondary outcomes, including Movement Disorder Society-sponsored revision of the Unified PD Rating Scale (MDS-UPDRS) Part III, were assessed in the ON-medication state at baseline and week 13.

**Findings:** Fifty participants were randomised to sham stimulation (Group A; n=25) or active CUE1+ stimulation (Group B; n=25). Median age was 70.0 years (IQR 65.0-75.5) in Group A and 68.0 years (62.0-75.0) in Group B. Group A included 14 (56%) men; Group B, 13 (52%) men. Four participants (8.0%) discontinued: three (6.3%) from Group A, one (2.0%) from Group B. Compliance to allocated intervention was equally excellent in both groups. Mild, transient skin irritation occurred in two participants (4.2%). MDS-UPDRS Part III scores improved by –4.5 points (95%CI: –8.7, –0.4; p=0.043), in Group A and –15.6 points (95%CI: – 20.3, –10.9; p<0.0001) in Group B, with a between-group difference of 11.08 points (95%CI: 4.85-17.31; p=0.002).

**Interpretation:** CUE1+ is a safe and well-tolerated non-invasive device that improves motor and non-motor outcomes in PD. These findings suggest a potential therapeutic benefit and support further evaluation in large-scale RCTs and consideration for integration into health care pathways.

**Funding:** UKRI Knowledge Transfer Partnership, 2021-2022, round 4 and Charco Neurotech Ltd.

## Introduction

Parkinson’s disease (PD) is a progressive neurodegenerative disorder affecting over 12 million people worldwide^1^. It is characterised by cardinal motor signs including bradykinesia, rigidity, and rest tremor, alongside axial features such as freezing of gait (FOG) and postural instability^2^. Symptomatic management remains the mainstay of treatment, with dopaminergic therapies, particularly levodopa, serving as the gold standard. While pharmacological agents effectively mitigate symptoms at the beginning of the disease course, their long-term use is associated with significant challenges, including motor fluctuations, dyskinesias, and diminished efficacy against axial symptoms^3,4^. As the demographic shift towards older populations continues, the prevalence, clinical complexity, and personal and public burden of PD are anticipated to rise steeply^1^, posing urgent challenges for healthcare systems and service planning^5^.

One major gap in current treatment lies in managing “OFF” periods, times when medication fails to control motor symptoms adequately^3,4^. These fluctuations often become more frequent and unpredictable as the disease progresses, leading to reduced health-related quality of life (QoL) and increased caregiver burden^1,5^. Although advanced therapies such as deep brain stimulation (DBS) and continuous infusion therapies are available in many countries, they are invasive, expensive, and not suitable for all patients^6,7^. Additionally, a wide array of prodromal, non-motor symptoms including hyposmia, depression, anxiety, cognitive dysfunction, REM sleep behaviour disorder, and autonomic dysfunction that may precede and progress alongside motor signs^2,8^. These non-motor features, are frequently under-recognised, often inadequately addressed by pharmacotherapy, substantially diminish QoL, and drive caregiver burden and healthcare demand^7^.

These challenges have led to growing interest in non-pharmacological strategies that target motor control and functional mobility without introducing additional systemic side effects. Among these, neuromodulatory wearables represent a promising frontier. The CUE1+ device, a non-invasive vibrotactile stimulator designed to be worn externally, aims to provide real-time sensory cueing with vibrotactile stimulation to improve motor function in people with PD^9,10^. Based on the principles of sensory-motor integration and cueing theory, such devices may help mitigate FOG, bradykinesia, and akinesia through peripheral stimulation and entrainment^9,11,12,13–15^.

Rhythmic sensory cues whether auditory, visual, or tactile may activate alternative neural pathways, compensating for dysfunctional basal ganglia circuits in PD^12,16^. By introducing external sensory input, vibrotactile cueing devices like CUE1+ may offer a safe, accessible, and user-controlled method to support mobility and independence in everyday settings^9^.

High-quality evidence for wearable vibrotactile cueing devices remains limited^11,12^, highlighting the need for blinded randomised controlled trials (RCTs) to confirm their benefits and assess long-term use. This double-blind RCT aimed to evaluate the usability, and safety/tolerability of CUE1+. Secondary exploratory outcomes included motor function improvement compared to sham device intervention, as well as gait, balance, falls, non-motor symptoms, and QoL. The trial also aimed to inform future RCT design and align with NICE and NHS England guidance^17,18^, which emphasise real-world, patient-centred studies. Interest in discreet wearable technologies is high among patients^19^. Based on prior findings^9^, we hypothesised successful recruitment, high adherence, minimal adverse events, and greater improvements in the active CUE1+ group.

## Methods

### Study design

This was a 12-week, multi-centre, double-blind, pilot RCT conducted at the Centre for Preventive Neurology, Queen Mary University of London (QMUL), in collaboration with Barts Health NHS Trust and Homerton Healthcare NHS Foundation Trust. The trial was approved by the London-Dulwich Research Ethics Committee (reference: 23/PR/1526), was registered prospectively with ClinicalTrials.gov (NCT06174948), and adhered to the principles outlined in the Declaration of Helsinki. An independent Patient and Public Involvement and Engagement advisory group contributed to the review of all study documentation and monitored trial progress. The protocol and statistical analysis plan were approved before the first participant was enrolled. The study protocol is available in the public domain^10^.

### Participants

Participants were identified from the Neurology Department at Barts Health NHS Trust and the Department of Care of the Elderly at Homerton Healthcare NHS Foundation Trust. All study assessments were conducted at the Centre for Preventive Neurology, QMUL. Written informed consent was obtained from all participants before any study-related procedures were undertaken.

Eligible participants met the following inclusion criteria: a clinical diagnosis of PD according to the Movement Disorder Society criteria^20^, age≥18 years, capacity and willingness to participate, and provision of written informed consent following review of the participant information sheet. Exclusion criteria included the presence of any medical or neurological conditions other than PD that could interfere with movement, balance, or independent participation in the trial. These included atypical parkinsonian syndromes, osteoarticular disorders, significant visual problems, audio-vestibular impairments, and a clinical diagnosis of dementia or Alzheimer’s disease. Participants were also excluded if they were receiving any non-standard therapeutic, cueing, or vibrotactile stimulation interventions, or if their Parkinson’s medication had not been stable for at least 3 months prior to enrolment. Additional exclusion criteria comprised the presence of a) implanted metallic or electronic devices such for DBS; b) known hypersensitivity to vibrotactile stimulation; and/or c) dermatological conditions or open wounds at or near the intended site of device application (i.e., the sternum).

Demographic and clinical data were collected at baseline, including age, sex, ethnicity, disease duration, disease severity (Hoehn and Yahr staging)^21^, and cognitive status as measured by the Montreal Cognitive Assessment (MoCA)^22^. A MoCA score ≥26/30 was interpreted as consistent with normal cognition. Sex and ethnicity were self-reported according to categories based on the UK Office for National Statistics^23^ classification. Participants could choose not to disclose this information. Ethnicity data were collected solely for descriptive statistics.

### Randomisation and masking

At the time of protocol publication^10^, treatment allocation was masked. After trial completion, Group A was unmasked as the sham intervention and Group B as the active non-invasive vibrotactile device. Masking of participants, investigators and assessor was maintained throughout the study until completion of data collection and analysis.

Randomisation was done centrally using a secure, web-based system (www.sealedenvelope.com), employing permuted blocks of varying sizes (2, 4, 6) to ensure allocation concealment and minimise predictability (Appendix 1). Multiple pre-labelled randomisation sequences (1-4) and block identifiers (1-14) were used to track allocation without compromising blinding. One researcher (CS), not involved in assessments or analysis, managed the randomisation process and held exclusive access to allocation codes. These codes were securely stored and inaccessible to other team members. Outcome assessments were done by VA, and data were analysed by VA and JPB, both masked to group allocation throughout. No emergency unblinding occurred or was anticipated, owing to the non-invasive nature of the intervention and the absence of adverse events in the feasibility study^9^.

### Procedures

Participants assigned to the experimental group (Group B) used the CUE1+ device for 12 weeks (intervention weeks 1-12). CUE1+ is a non-invasive, MHRA-registered medical device developed by Charco Neurotech Ltd (Appendix 2). It delivers combined tactile cueing and vibrotactile stimulation and is worn on the sternum using a dermatologically approved adhesive patch. Participants were instructed to wear the device daily for 8 hours, starting each day from the morning, beginning one hour after taking their routine dopaminergic medication. Stimulation was delivered at 80% of the device’s maximum vibration strength, with pulse and rest intervals of 800 ms each, based on effective parameters reported in previous work^9,10^. Participants in the control group (Group A) received an identical sham device, visually and functionally indistinguishable from the active version but with vibrotactile output disabled (0% vibration strength; 0 ms pulse and rest duration). To maintain blinding, participants were informed that stimulation might be subtle or imperceptible. The usage protocol was identical that of Group B.

Both groups received training and written instructions, including a support booklet detailing device operation, patch application, and charging. Additional support was provided as needed during the trial. Supplementary instructions were made available and are accessible with the published protocol^10^.

Participants continued their usual antiparkinsonian medications throughout the trial. Any necessary medication changes led to participant withdrawal, at which point data collection ceased. Consent was obtained to retain data collected up to withdrawal. Participants could also withdraw at any time, with previously collected data retained if they agreed. All protocol deviations and violations were reported to the study sponsor. Following unblinding, Group A participants were informed of their allocation, asked to return the sham device, and offered an active CUE1+ for use outside the trial. All participants were offered the CUE1+ upon trial completion, irrespective of group

### Outcomes

The primary outcome was CUE1+ usability at home over 12 weeks, including recruitment rate, compliance with intervention, dropout rates, and safety/tolerability. Site-level usability included engagement and recruitment performance. Protocol compliance was monitored through adherence to core study procedures, including timely delivery of interventions, consistency in outcome assessments, and data collection completeness. Investigators also recorded the proportion of people screened who met inclusion criteria, were approached for participation, consented, and were ultimately randomised.

Participant-level usability included compliance to the allocated intervention, retention through the 12-week follow-up, and completeness of baseline and outcome data. Compliance with the assigned intervention across the 12-week period was assessed using two self-report measures. Participants were first asked to indicate their usage frequency of the CUE1+ device on a 5-point Likert scale ranging from 0 (did not use it at all) to 4 (used it daily as advised, i.e., 8+ hours/day). Additionally, they were asked to provide a subjective rating of their overall compliance on a continuous scale from 0 (not compliant at all) to 10 (completely compliant). Safety and tolerability were monitored throughout, including any clinical events and adverse events. Device malfunctions in both active and sham groups were logged. In the end of the study, all participants completed a satisfaction questionnaire on their experience with CUE1+, as published with the trial protocol^10^. The questionnaire utilised a 5-point Likert scale, ranging from 0 (not at all) to 4 (extremely), to assess various aspects of participant satisfaction. Scores for all endpoints were subsequently compared between groups.

Clinical visits were conducted at baseline (week 0) and follow-up (week 13), each lasting approximately half a day. These visits included motor assessment and completion of self-reported outcome questionnaires. To mitigate fatigue-related bias, the order of assessments was randomised, and rest breaks were offered. The full assessment schedule is available in the published protocol^10^.

Secondary exploratory outcomes included clinical efficacy measures. Details on all clinical efficacy measures, scoring procedures, minimal clinically important differences (MCIDs) and additional reference list are provided (Appendix 3). These outcomes were evaluated to inform the design of a future definitive RCT. Motor assessments included the Movement Disorder Society-sponsored revision of the Unified Parkinson’s Disease Rating Scale (MDS-UPDRS) Part III^24^, Functional Gait Assessment^25^, Timed Up and Go (TUG)^26^, TUG with dual-task using serial sevens, and the two keyboard-based tapping tests to measure hand dexterity and movement (e.g., kinesia), Bradykinesia Akinesia INcoordination (BRAIN-Kinesia Score; KS)^27^ and Digital Finger Tapping (DFT-KS)^28^ tests for the most affected side. Optional video recordings of the MDS-UPRDS Part III assessments were retained for educational purposes to QMUL, with participant consent. All motor testing was performed in the ON-medication state (45-60 minutes after dopaminergic therapy). Participants stopped wearing the device just before attending the follow-up assessment to maintain blinding of the assessor.

Another secondary exploratory measure was the Parkinson’s KinetiGraph (PKG)^29^ wristwatch (Appendix 3). Participants wore the PKG wristwatch continuously on their most affected side for 13 weeks, beginning on the day of the baseline assessment (week 0) and concluding after the week 13 follow-up. They only removed the PKG when this needed to be charged for approximately two hours every 6-7days. The main data points collected from the PKG system are also included in the published protocol^10^.

Patient-reported outcomes covered both motor and non-motor domains and included the MDS-UPDRS Parts I, II, and IV^24^, Activities-specific Balance Confidence Scale (ABC)^30^, Pittsburgh Sleep Quality Index (PSQI)^31^, Parkinson’s Disease Questionnaire–39 (PDQ-39)^32^, Hospital Anxiety and Depression Scale (HADS)^33^, Fatigue Severity Scale (FSS)^34^, and Patient Global Impression of Change^35^.

## Statistical analysis

Statistical analyses were conducted using IBM SPSS Statistics (version 29; IBM Corp., Armonk, NY, USA). Descriptive statistics were used to summarise baseline characteristics and outcome data. For continuous variables, normally distributed data are presented as mean (standard deviation; SD), and non-normally distributed data as median (interquartile range; IQR). Categorical variables are shown as absolute (number; n) and relative (percentage; %) values. Normality of distributions was assessed using the Shapiro-Wilk test, supported by histogram and Q–Q plot inspection. Ninety-five per cent confidence interval (95% CI) are reported where appropriate to indicate precision.

Between-group comparisons at baseline and follow-up were conducted using independent samples t tests for normally distributed variables, and Mann-Whitney U tests for non-normally distributed variables. Within-group comparisons were assessed using paired t-tests (normal distribution) or Wilcoxon signed-rank tests (non-normal distribution). When one timepoint was normally distributed and the other was not, non-parametric methods were used to preserve validity. An intention-to-treat (ITT) approach was used for all clinical efficacy outcomes. Data from participants who discontinued the intervention or deviated from the protocol were retained in the analysis unless consent was withdrawn. To enhance clinical interpretability, results were also assessed using established MCIDs for outcomes related to PD, where available. MCIDs provide a more patient-centred benchmark than purely statistical metrics such as minimal detectable change (MDC)^36^. Bonferroni correction was considered for multiple comparisons of secondary effectiveness outcomes (e.g., p<0.004 for 14 outcomes).

As a pilot RCT, no priori power calculation was performed. The sample size was guided by Sim and Julius^37^, who recommend 20-75 participants per arm to detect moderate to large effects. Drawing from feasibility data, including a previous single-arm study^9^ involving ten participants, the target sample was initially set at 20 per group. To account for an anticipated dropout rate of up to 25% given the use of a sham comparator, the final recruitment goal was increased to 25 participants per group. Post hoc sample size and power calculations for a future definitive RCT were conducted. Parametric or non-parametric methods were used depending on data distribution, accounting for dropout, and targeting 80% power at alpha (a) equal 0.05.

## Results

The trial was completed as planned, without delays or protocol amendments. As outlined in the protocol^10^, several exploratory secondary outcomes were pre-specified, though PKG data were not included. During data analysis, PKG monitors showed substantial variability in data completeness and standardisation. To maintain scientific validity follow-up PKG data (weeks 1-13) were excluded. Baseline PKG data (week 0), recorded before intervention, are included for between-group comparison of real-world activity. This approach aligns with best practice guidelines for reporting and data integrity^38^.

Between September 1, 2024, and February 28, 2025, a total of 58 individuals were screened for eligibility: 13 (22.4%) at Homerton University Hospital and 45 (77.6%) at Barts Health NHS Trust. Of those screened, 5 (8.6%) did not meet inclusion criteria, and 3 (5.2%) declined participation due to personal reasons (Figure 1). The remaining 50 were randomly assigned to the control group (Group A) or the intervention group (Group B). Of those randomised, 7 (14.0%) were recruited from Homerton University Hospital and 43 (86.0%) from Barts Health NHS Trust. Baseline characteristics of all participants are shown in Table 1. Seven (30%) of participants in Group A and 11 (44%) in Group B were from non-white ethnic origin.

**Figure 1:**
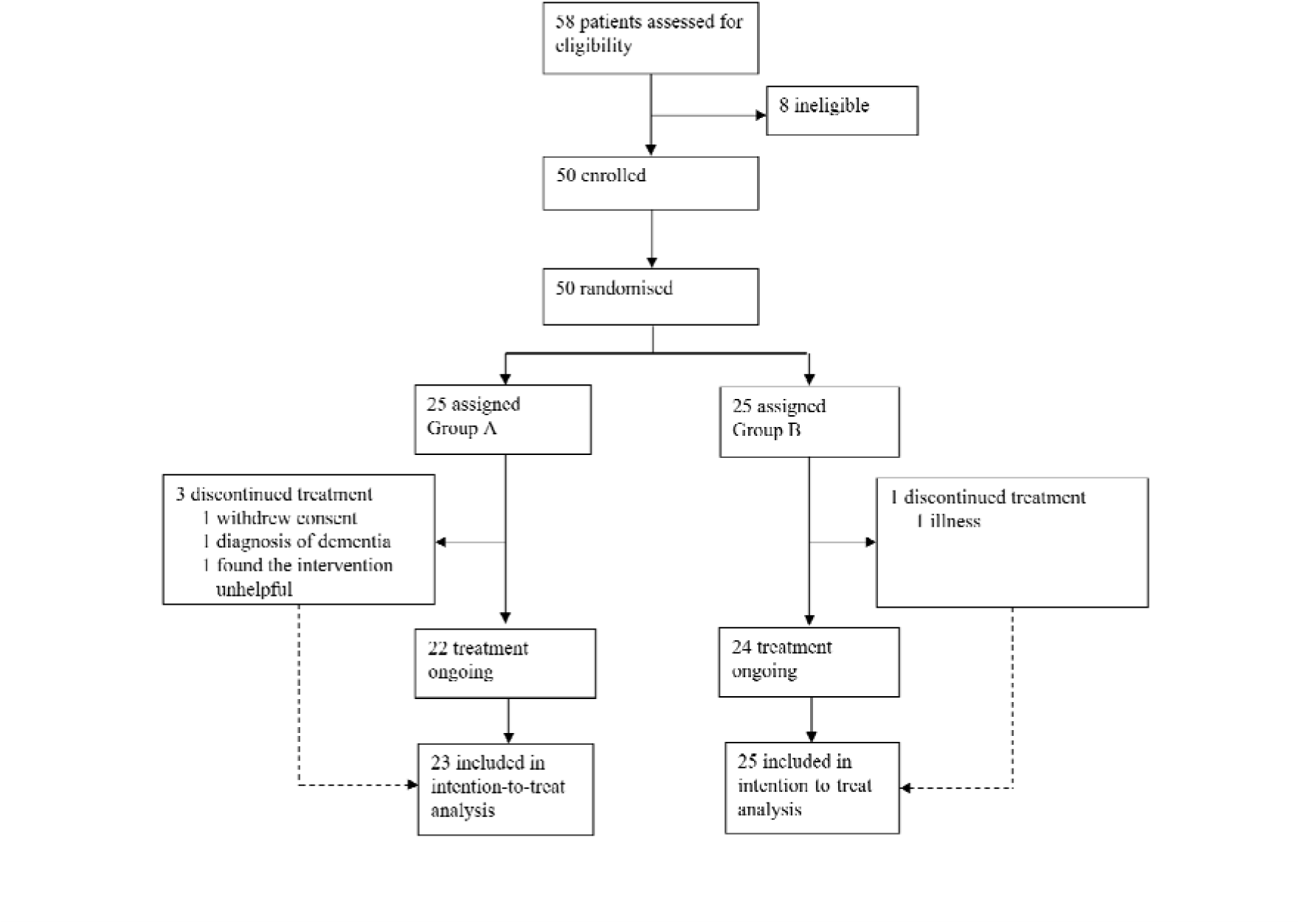
Trial profile. Group A involved participants who received sham device intervention while Group B involved participants who received active device intervention.

**Table 1:** Baseline characteristics of the intention-to-treat population.

|  |  | Sham device group<br>(n = 23)† | Active device group<br>(n = 25)† |
| --- | --- | --- | --- |
| Age, years |  | 70.0 (65.0, 75.5) | 68.0 (62.0, 75.0) |
| Sex |  |  |  |
|  | Male | 14 (56%) | 13 (52%) |
|  | Female | 9 (36%) | 12 (48%) |
|  | Non-binary | 0 (0%) | 0 (0%) |
|  | Prefer not to say | 0 (0%) | 0 (0%) |
| Ethnic origin |  |  |  |
|  | White | 16 (70%) | 14 (56%) |
|  | Black | 0 (0%) | 3 (12%) |
|  | Asian | 7 (30%) | 8 (32%) |
|  | Mixed | 0 (0%) | 0 (0%) |
|  | Other | 0 (0%) | 0 (0%) |
| Symptom duration, years |  | 5.0 (2.5, 7.0) | 5.0 (2.0, 8.0) |
| Symptom severity | Hoehn & Yahr | 3.0 (2.5, 3.0) | 3.0 (2.0, 3.0) |
| Most affected side by<br>Parkinson's as per<br>clinical assessment |  |  |  |
|  | Right | 14 (56.0%) | 11 (44.0%) |
|  | Left | 10 (43.5%) | 13 (56.5%) |
| Montreal Cognitive<br>Assessment Tool |  | 26.0 (24.5, 28.0) | 25.0 (23.0, 27.0) |

Data are n (%), or median (IQR). N= number.
†Sham device group is referred as Group A in the manuscript while active device group as Group B.

Recruitment rate was considered high (86%) consistent with our previous clinical study^9^, which likely reflects the non-invasive nature of the intervention and strong site engagement. Four participants (8.0%) discontinued the trial, 3 (6.3%) from Group A and 1 (2.0%) from Group B. The overall dropout rate was low and within the expected range for a study of this design and duration.

The intervention was well tolerated across both groups, with minimal adverse events. Mild, transient skin irritation at the adhesive patch site occurred in two participants (4.2%), one in each group, and resolved without treatment. One Group B participant (2.1%) reported dizziness during use at the pre-set vibration intensity; however, this coincided with generalised dizziness unrelated to the device, and the participant withdrew due to an intercurrent illness. After the trial, they resumed use of the CUE1+ independently and increased vibration intensity from 50% to 70% via the CUE App (not used during the trial), without recurrence of dizziness. No participants reported difficulty with patch application or removal. A single technical malfunction occurred in Group B and was promptly resolved by an unblinded researcher (CS) in coordination with Charco Neurotech Ltd. A replacement device was delivered within days, and no further technical issues were reported. Overall, the intervention demonstrated a favourable safety and usability profile.

Participant-level usability and compliance were excellent with no notable between-group difference (Table 2). Participants in Group B found the CUE1+ more helpful in managing their symptoms compared with those in Group A [-1.1 (−1.7, –0.5), p=0.001] (Table 3). Group B expressed a higher likelihood of continuing device use beyond the trial period [1.0 (0.5, 1.0), p=0.004]. However, there was no difference in whether participants in groups A and B would recommend the intervention to others with PD [0.0 (0.0, 1.0), p=0.336].

**Table 2.** Participant-level usability and compliance to intervention.

| Compliance |  | Sham device group†††<br>(n = 23) | Active device group†††<br>(n = 25) |
| --- | --- | --- | --- |
| How compliant were you with the use of the CUE1+ device during the study? † | 4-Used it daily as advised (8+ hours/day, nearly every day) | 22 (95.7%) | 25 (100.0%) |
|  | 3-Used it most days but not always 8 hours | 0 (0.0%) | 0 (0.0%) |
|  | 2-Used it several times a week but inconsistently | 1 (4.3%) | 0 (0.0%) |
|  | 1-Rarely used it | 0 (0.0%) | 0 (0.0%) |
|  | 0-Did not use it at all | 0 (0.0%) | 0 (0.0%) |
| Could you please give a score from 0 to 10 for how compliant you feel you were to the CUE1+ in the study? †† | 10: Fully compliant- Used the device daily as advised (i.e., ≥8 hours/day), with no perceived deviation from the recommended usage. | 21 (91.3%) | 21 (84.0%) |
|  | 7-9: High compliance- Generally consistent use, with only occasional lapses in either frequency or duration. | 1 (4.3%) | 4 (16.0%) |
|  | 4-6: Moderate compliance- Some degree of regular use, but with notable deviations from the daily, full-duration recommendation. | 1 (4.3%) | 0 (0.0%) |
|  | 1-3: Very low compliance- Minimal or sporadic use, well below the advised frequency or duration. | 0 (0.0%) | 0 (0.0%) |
|  | 0: Not compliant at all – No effort made or complete failure to use the device throughout the study period. | 0 (0.0%) | 0 (0.0%) |

Data are n (%). N= number.
† Indicates items scored on a 5-point Likert scale ranging from 0 to 4.
†† Indicates items scored on a Visual Analogue Scale ranging from 0 to 10.
††† Sham device group is referred in the main manuscript as Group A while active device group as Group B.

**Table 3.**
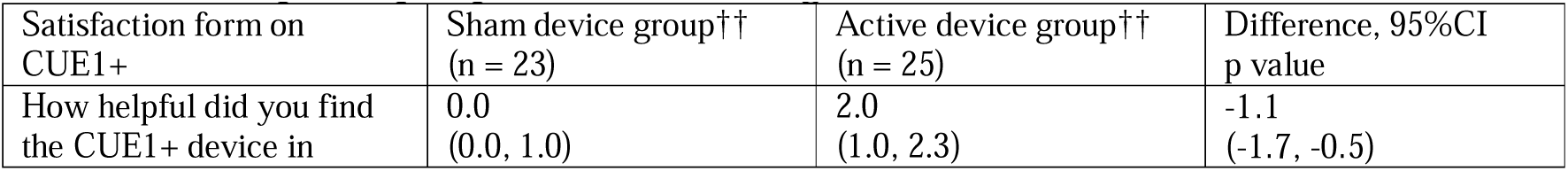
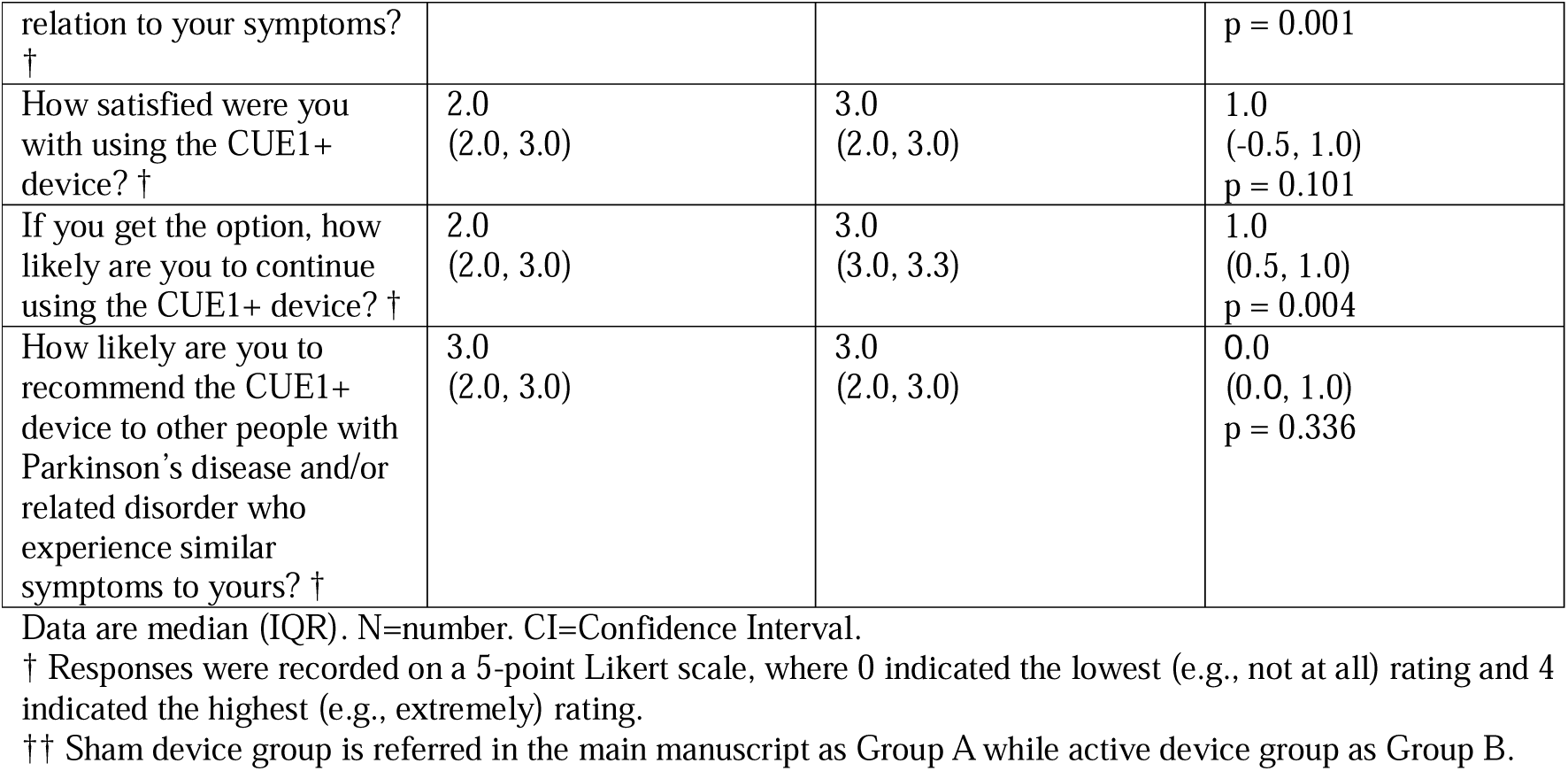
Participants’ perspective on receiving CUE1+ intervention.

At baseline, there were not significant between-group differences for secondary effectiveness outcomes (Appendices 4 and 5). At follow-up, significant between-group improvements were noted on MDS-UPDRS Part III [11.1 (4.9, 17.3; p=0.002] (Figure 2) and PDQ-39 [Summary Index; SI: –7.6 (−12.3, –3.0), p=0.003] in favour of Group B (Appendix 6). The median PGIC “change since care” score at follow-up was 2.0 in Group A (1.0, 3.5) and 4.0 in Group B (2.0, 5.0), with a trend toward better ratings in Group B (p=0.051). The PGIC “degree of change” score was 5.0 in Group A (4.0, 5.0) and 3.0 in Group B (3.0, 4.0), showing a more notable between-group difference in favour of Group B [-2.0 (−2.0, –0.5), p=0.012].

**Figure 2:**
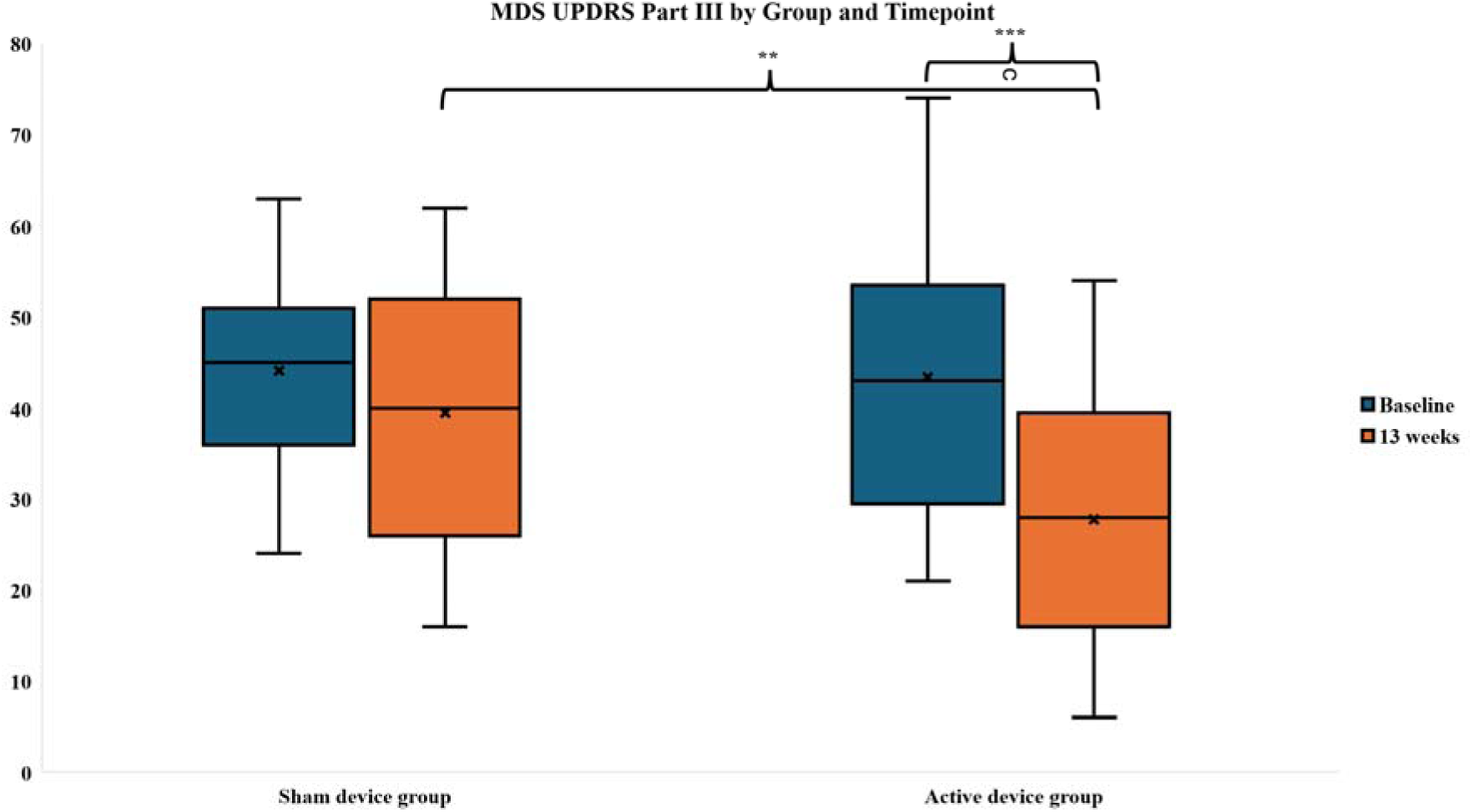
Box plot (median, confidence interval, 1st and 3rd quartiles, maximum and minimum) showing the results of MDS-UPDRS Part III scores at baseline and week 13 for sham and active device group. The sham device group showed a mean change of –4.5 points (95% CI: –8.7, –0.4; p = 0.043), while the active device group had a mean change of –15.6 points (95% CI: –20.3, –10.9; p < 0.0001). The between-group difference in change was 11.1 points (95% CI: 4.9, 17.3; p = 0.002). After applying the Bonferroni correction, the threshold for statistical significance was adjusted from p < 0.05 to p < 0.004. **p<0.01, ***p<0.001. Sham device group is referred as Group A in the main manuscript while active device group as Group B.

Participants wearing the active device (Group B) showed a notably greater within-group improvement on objective motor outcomes compared to those who received the sham device (Group A). This included significantly greater improvements on the MDS-UPDRS Part III [Group B: –15.6 (−20.3, –10.9), p<0.0001; Group A: –4.5 (−8.7, –0.4), p=0.043] (Figure 2), and on the BRAIN KS test [Group B: 3.0 (0.0, 12.0), p=0.007; Group A: 3.0 (0.0, 7.0), p=0.056], although the within-group changes in the latter were not statistically significant. Group B also showed more notable improvements on the FGA [Group B: 3.0 (2.0, 4.0), p<0.0001; Group A: 1.2 (0.0, 2.5), p=0.053] (Appendix 7).

These motor improvements were further supported by participant self-reported outcomes on motor symptoms, specifically MDS-UPDRS Part II [Group B: –4.0 (−7.0, –1.0), p<0.0001; Group A: 0.0 (3.0, 0.0), p=0.980], as well as Part I [Group B: –4.0 (7.0, 2.0), p<0.0001; Group A: 0.0 (3.0, 0.0), p=0.410] and Part IV [Group B: –4.0 (5.0, –3.0), p<0.0001; Group A: 0.0 (0.0, 1.0), p=0.850].

In addition, improvements were observed in self-reported outcomes on non-motor symptoms, including the PSQI [Group B: –2.0 (−3.0, –1.0), p<0.0001; Group A: –0.7 (−2.2, 0.9), p=0.401], PDQ-39 Summary Index [Group B: –7.5 (−11.5, –3.4), p<0.0001; Group A: 0.1 (−2.9, 3.1), p=0.925], and the PDQ-39 Activities of Daily Living component [Group B: –12.5 (−12.5, – 4.2), p<0.0001; Group A: –4.2 (−8.3, 0.0), p=0.073].

A post hoc power analysis was conducted based on changes in MDS-UPDRS Part III, the gold standard for motor assessment in PD^24^. Participants in Group B demonstrated a mean improvement of 15.6 points (SD=13.3), compared to a 4.5-point improvement (SD=13.2) in Group A, yielding a mean between-group difference of 11.1 points and a standardised effect size (Cohen’s d) of 0.88. Based on this effect size, a future two-arm, fully powered RCT would require 33 participants per group to achieve 80% power and 52 participants per group to achieve 95% power, assuming a two-tailed alpha of 0.05. Accounting for the observed dropout rate of 8%, the adjusted sample size estimates increase to 36 and 57 participants per group for 80% and 95% power, respectively. However, accounting for a projected dropout rate of 25% due to a sham intervention involvement, the adjusted sample size estimates would increase to 44 and 70 participants per group for 80% and 95% power, respectively. These findings suggest that a fully powered RCT is feasible and could be conducted with a relatively modest sample size.

## Discussion

This double-blind, multicentre double-blind RCT provides robust preliminary evidence supporting real-world usability, safety/tolerability and clinical efficacy of the CUE1+ device in management of PD. Compared with the sham control, participants who received the active device experienced greater clinically meaningful improvements across multiple domains. These included motor severity and complications of treatment, functional gait, falls risk, non-motor symptoms such as sleep quality, self-reported impression of change, and overall QoL over 12 weeks. These benefits were achieved with high recruitment rate and compliance to intervention, minimal adverse effects, and positive participant experience, factors which are important when considering integration into routine care.

Although 43.5% of participants using the sham device met the MCID threshold on the MDS-UPDRS Part III, consistent with known placebo responses in PD when exposed to interventions^14^, the significantly greater proportion achieving this in the active device group (84%) supports a genuine therapeutic effect. In our study, we used –3.25 points as the MCID cutoff, based on Horváth et al.^39^ who employed robust anchor-based methods in a large longitudinal cohort. In contrast, Sánchez-Ferro et al.^40^ proposed a more conservative ±5-point threshold using the original UPDRS-III in a pragmatic setting. The discrepancy likely reflects differences in methodology, population characteristics, and particularly, the use of the revised MDS-UPDRS versus the original UPDRS. Notably, the threshold in the study by Sánchez-Ferro et al.^40^ applies to UPDRS-III, while the estimate in study conducted by Horváth et al.^39^ concerns MDS-UPDRS Part III, which may be more sensitive to change as this was assessed by anchor_based methods^39^. Standardising MCID definitions across UPDRS versions remains critical for consistent interpretation and comparison.

Falls remain a major issue in PD, with 45% to 60% falling each year and two-thirds experiencing recurrent falls^41^. While this study indicated reduced fall risk (e.g., FGA) following the CUE1+ intervention, the MCID^25^ was achieved only by 20% of participants. Longer duration follow-up might be required to observe meaningful changes in more patients and to confirm sustained benefit. Future research should also examine how vibrotactile stimulation affects medication use in PD, particularly given observed improvements in motor complications assessed on MDS-UPDRS Part IV. An observational study showed levodopa doses remained stable in patients using vibrotactile devices, while increasing in others, suggesting better symptom control^15^, potentially also reducing falls risk.

Sleep disturbances are prevalent in PwP^42^ and were evident in this study, with participants exhibiting abnormal PSQI scores. Although scores improved following the intervention, they remained clinically abnormal, highlighting the potential need for longer or sleep-targeted adjunctive therapies. Given the bidirectional relationship between motor severity and sleep symptoms, addressing one may improve the other^42,43^. Future research should employ more sensitive sleep assessments and explore combined approaches, such as vibrotactile cueing with sleep interventions. Improvements in sleep and balance confidence may reflect not only motor gains but also vagally mediated effects on arousal, anxiety, and overall QoL^4^. The later might be improving also due to improvements in motor symptoms.

The findings from this study emerge at a critical time in UK policy, as NHS England and NICE seek to integrate evidence-based, patient-led digital health solutions into care pathways^17,18^. NICE 2023 request for evidence generation positions remote monitoring, wearable devices, and non-pharmacological strategies as key priorities for service innovation. The CUE1+ aligns directly with these aims. It has demonstrated applicability across motor and non-motor domains with no serious adverse events, making it a suitable candidate for broader implementation and NICE Medical Technologies Evaluation Programme (MTEP) review^44^. MTEP is recommended to support system-wide adoption, integration with NHS Digital frameworks. A targeted evaluation of cost per quality-adjusted life year (QALY) gained, drawing on improvements in motor function, falls prevention, and sleep, could justify scale-up. The pragmatic, home-based design and positive real-world usability profile of CUE1+ position it as a strong candidate for future commissioning decisions.

According to current treatment frameworks outlined in the updated NICE pathways for managing PD^45^, there may be a need for modalities and treatment options to be explored to support and enhance physiotherapy, occupational therapy, and speech and swallowing interventions, areas often limited by barriers such as cost, travel demands, and reduced patient motivation^43^. Future research should explore the implementation of vibrotactile devices designed for home use including CUE1+ across multiple care pathways. Furthermore, as CUE1+ shows potential in addressing postural instability, falls, and motor complications, this may position the device as a valuable adjunct in the management of complex, late-stage PD. Its use and contribution to reduce hospitalisation rates and caregiver burden, should be investigated in advanced care settings.

This RCT provides a foundation for a larger, multisite fully powered trial with a sample size estimation, and long-term follow-up to confirm efficacy and examine diversity in subgroup responses based on factors such as disease duration and severity, fall history, and cognitive status. Although not powered for economic evaluation, the large effect size (Cohen’s d=0.88) supports the need for future cost-utility analyses comparing CUE1+ with standard care, incorporating QALYs and NHS resource use to inform potential NICE Medical Technologies Guidance^44^. Future studies should also include head-to-head comparisons with other non-invasive neuromodulation devices (e.g., vibrotactile insoles, gloves, acoustic metronomes) to refine indications and positioning. Neuroimaging and electrophysiology could clarify the neural basis of clinical improvements, particularly in motor network reorganisation and sensorimotor integration^12^. Concurrently, wearable sensors such as accelerometers and fall detectors should continue to be used to collect real-world data which may help generate objective gait and posture data, enabling digital biomarker development. Real-world implementation will require research into clinician and patient uptake, digital inclusion, training needs, and equitable access. Co-design with people living with PD, caregivers, and healthcare professionals will be critical to ensure the device is acceptable, accessible, and seamlessly integrated into routine care.

This study has several limitations. The sample size, while sufficient for preliminary assessment, limits generalisability and prevents detailed subgroup analyses. Key questions remain about whether vibrotactile stimulation improves symptoms less responsive to medication, such as tremor and FOG, and whether benefits extend across diverse populations. The study was also not powered to assess economic outcomes or specific effects in underrepresented groups, including those with advanced PD, cognitive impairment, or non-English-speaking backgrounds. The 12-week follow-up period restricts conclusions about long-term efficacy and sustainability of benefits especially around falls and QoL. Fixed stimulation parameters were used throughout, meaning the potential advantages of personalised dosing were not assessed. Compliance and some outcomes were self-reported, introducing the possibility of bias despite supporting in-person assessments.

To conclude, the CUE1+ device appears to be a promising, non-invasive adjunct for managing motor and non-motor symptoms in PD, offering meaningful improvements across multiple domains of function and QoL. Its real-world usability, safety and tolerability suggests strong potential for integration into NHS care pathways. These findings support further assessment in a definitive RCT.

## Contributors

VA, AJN, and CS (along with Patient and Public Involvement and Engagement members) were involved in the conception, design and writing and editing of the study protocol. VA, CS and AJN were involved in the conception and editing of the protocol. VA, EB, EC, KCD, AZ, CQ, CB, TB, DG, AJN, and CS were involved in participants’ identification. JPB carried out the statistical analysis and reviewed the methodology and results sections of the manuscript. VA was involved in participants’ recruitment and assessments. CS was responsible for randomisation and blinding procedures. All authors reviewed and approved the final version of the manuscript prior submission. AJN and CS contributed equally to this manuscript as senior authors.

## Data sharing

Access to the dataset can be granted by contacting the corresponding author. Authors are committed to making the research data that supports this article available, wherever legally and ethically feasible. Therefore, data collected for the study, including deidentified individual participant data and a data dictionary defining each field in the set, will be made available to others. Additional, related documents can be also available upon request (e.g., study protocol, statistical analysis plan, informed consent form). This data sharing plan is in line with International Committee of Medical Journal Editors guidelines. The authors have updated the clinical trial registry to ensure compliance with the scientific peer-preview journals data sharing policy.

## Data Availability

https://clinicaltrials.gov/study/NCT06174948

## Acknowledgments

We would like to thank all participants for taking part in this study. We appreciate the efforts of all the authors for this article. This study is part of the project funded by Knowledge Transfer Partnership (KTP) United Kingdom 2021 to 2022, round 4, UKRI KTP (Innovate UK) and we would like to thank the organisation for the funding. We would like to thank Charco Neurotech Ltd for providing the CUE1+ devices and adhesives for the duration of the study, and for generously gifting CUE1+ devices along with a three-month supply of adhesives to all participants who wished to continue using the device post-study. We also gratefully acknowledge Charco Neurotech Ltd for covering the costs associated with the PKG wristwatches used in the study. Finally, we are grateful to the support that Ms Kira Rowsell has offered with communicating CUE1+ information at Homerton University Hospital.

## Appendices

## Appendix 1: Randomisation list

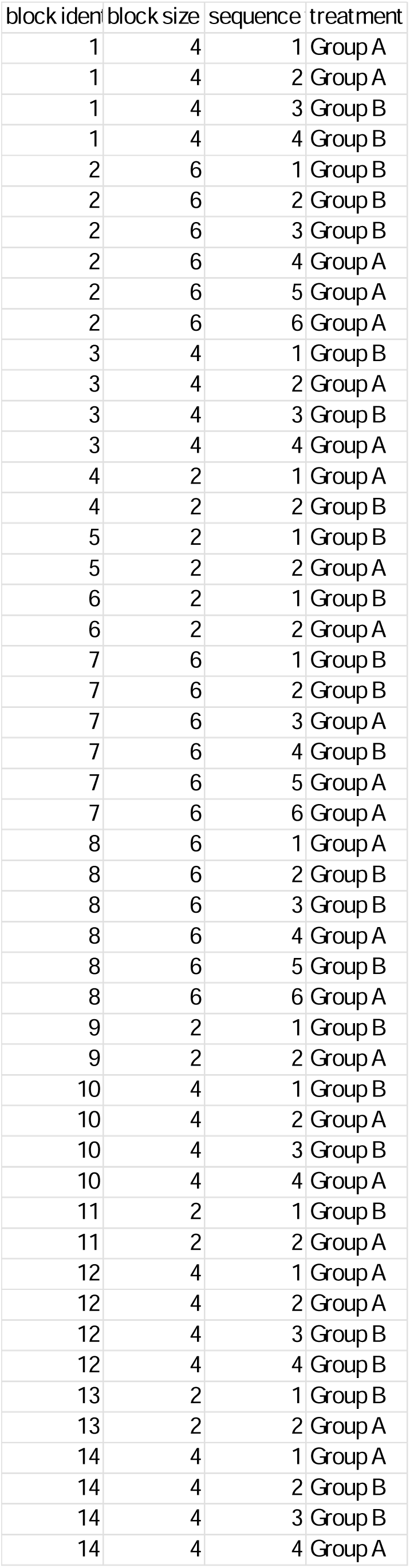

## Appendix 2: The CUE1+ device

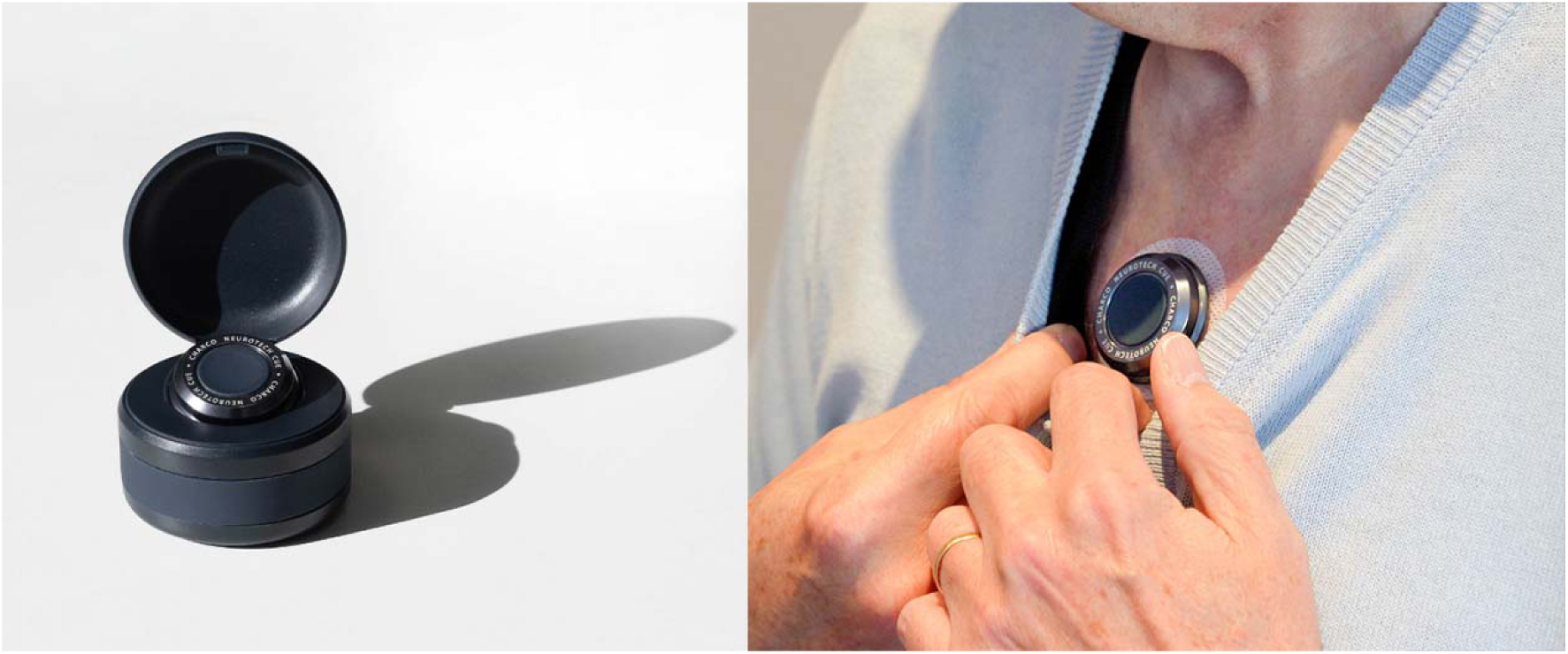

The CUE1+ is a non-invasive medical device designed to support the management of both motor and non-motor symptoms in people living with Parkinson’s. The device is compact and discreet (dimensions: 40 mm in diameter, 11 mm in height; weight: 17 g) and is affixed to the sternum using dermatologically tested adhesive patches. While it is water-resistant, it is not waterproof and should be removed prior to showering. When activated, the device delivers targeted vibrotactile stimulation via a silent motor, employing a proprietary waveform informed by extensive user feedback and clinical research. It integrates low-frequency auditory cueing with high-frequency vibrotactile stimulation, delivered in a structured pattern intended to optimise therapeutic benefit. A sham version of the CUE1+ is used as a control within the trial. It is externally identical to the active device, but its therapeutic functions have been fully disabled.

## Appendix 3: Description of motor and non-motor outcomes

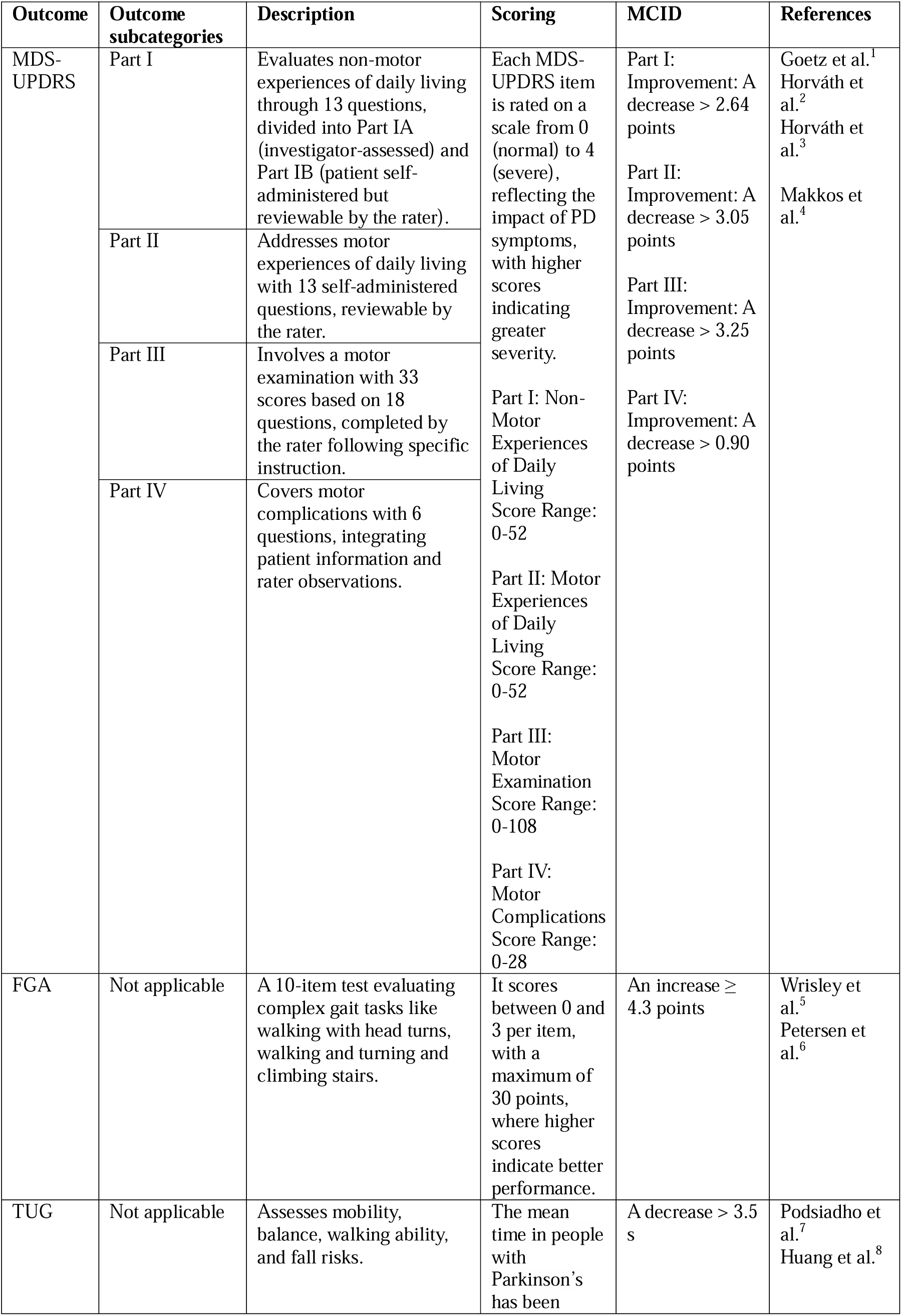
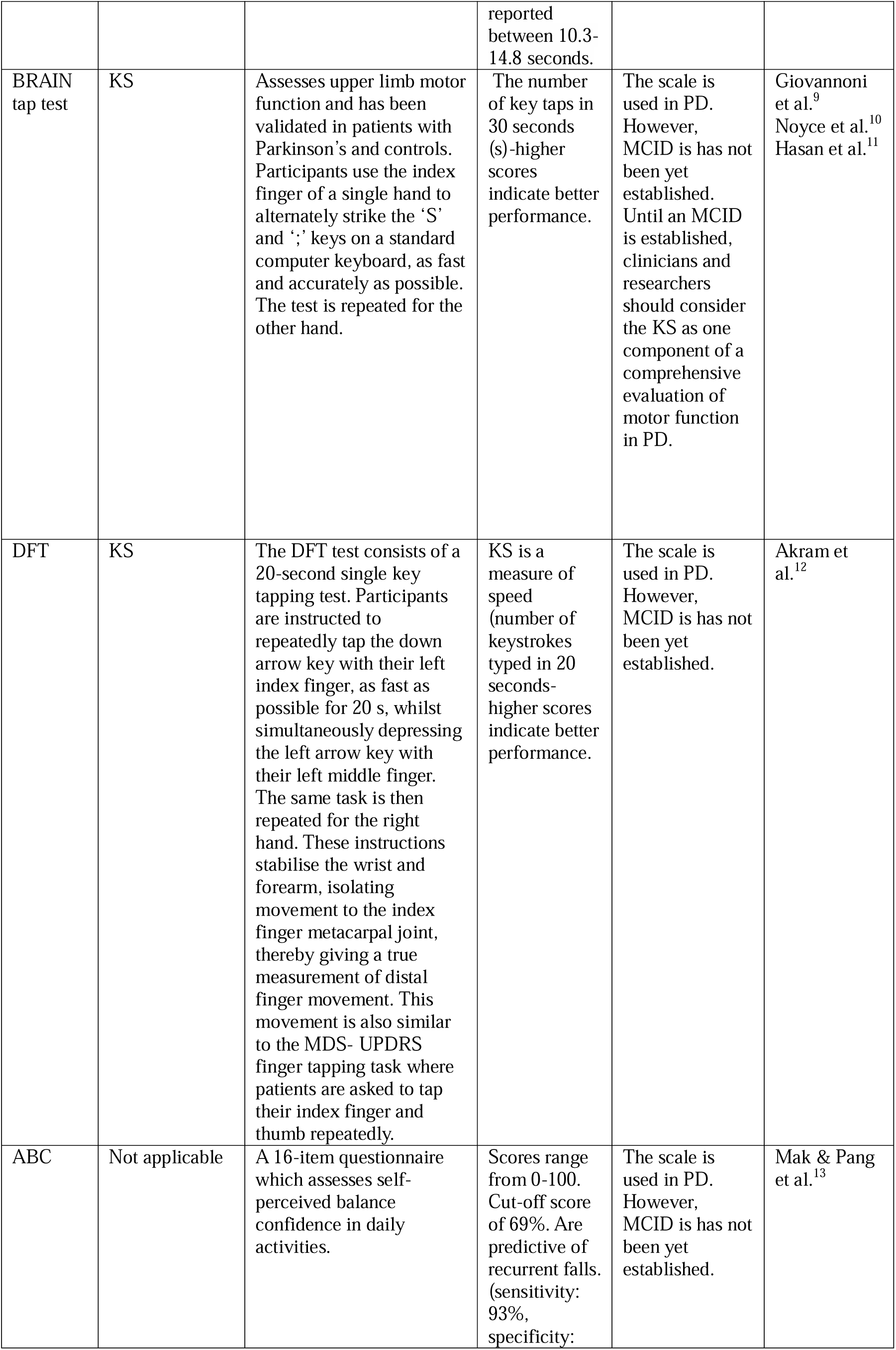
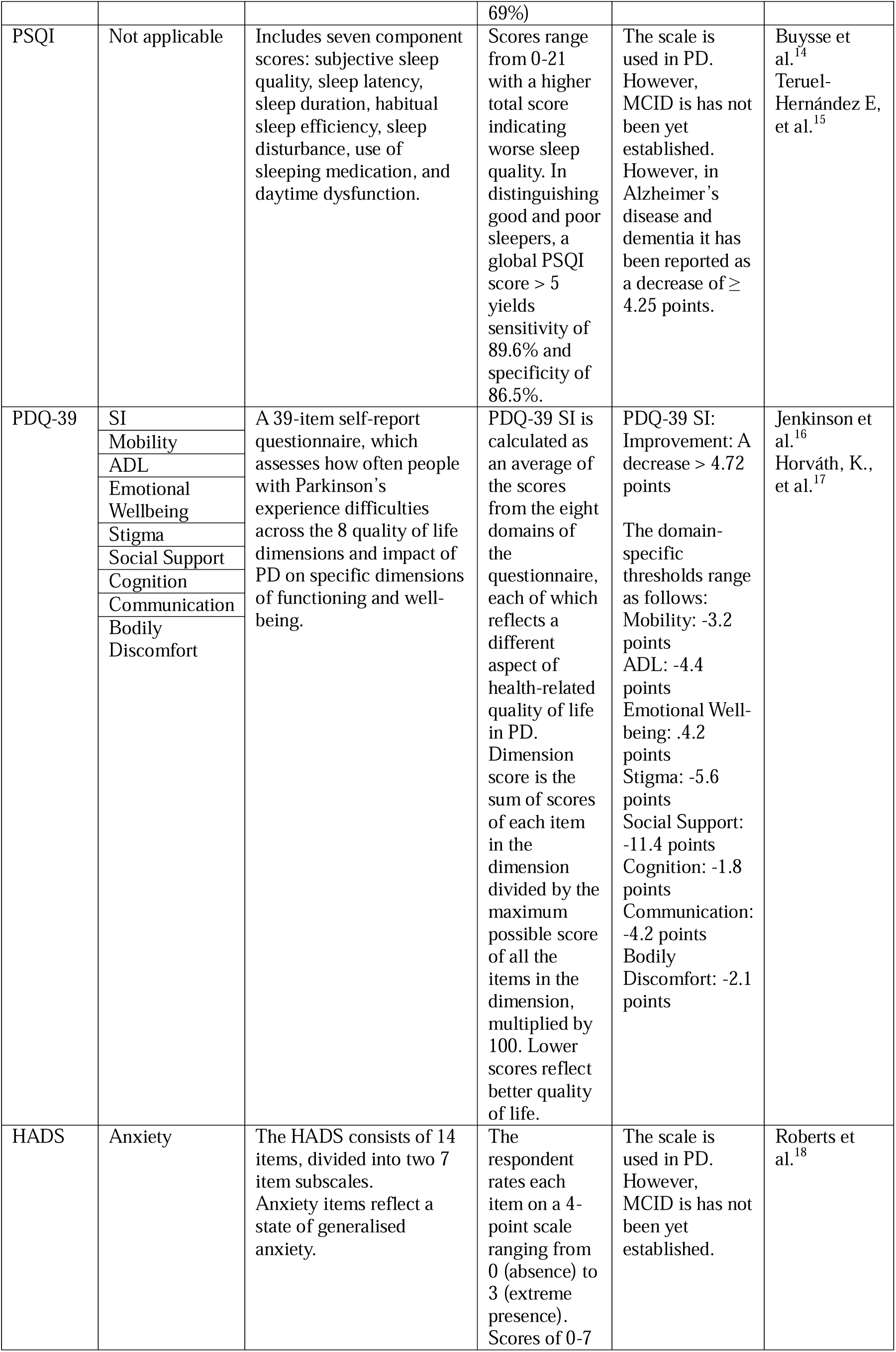
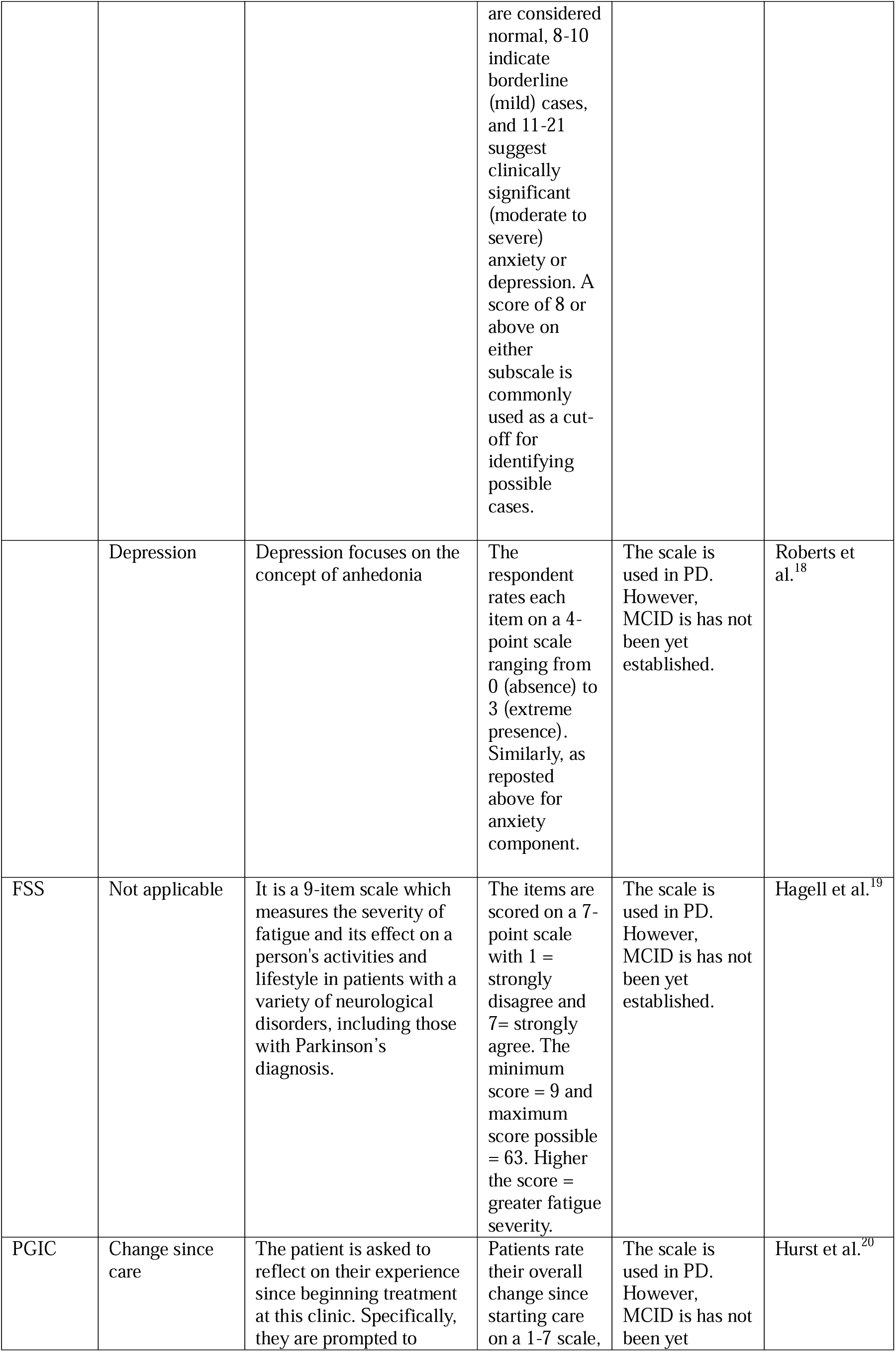
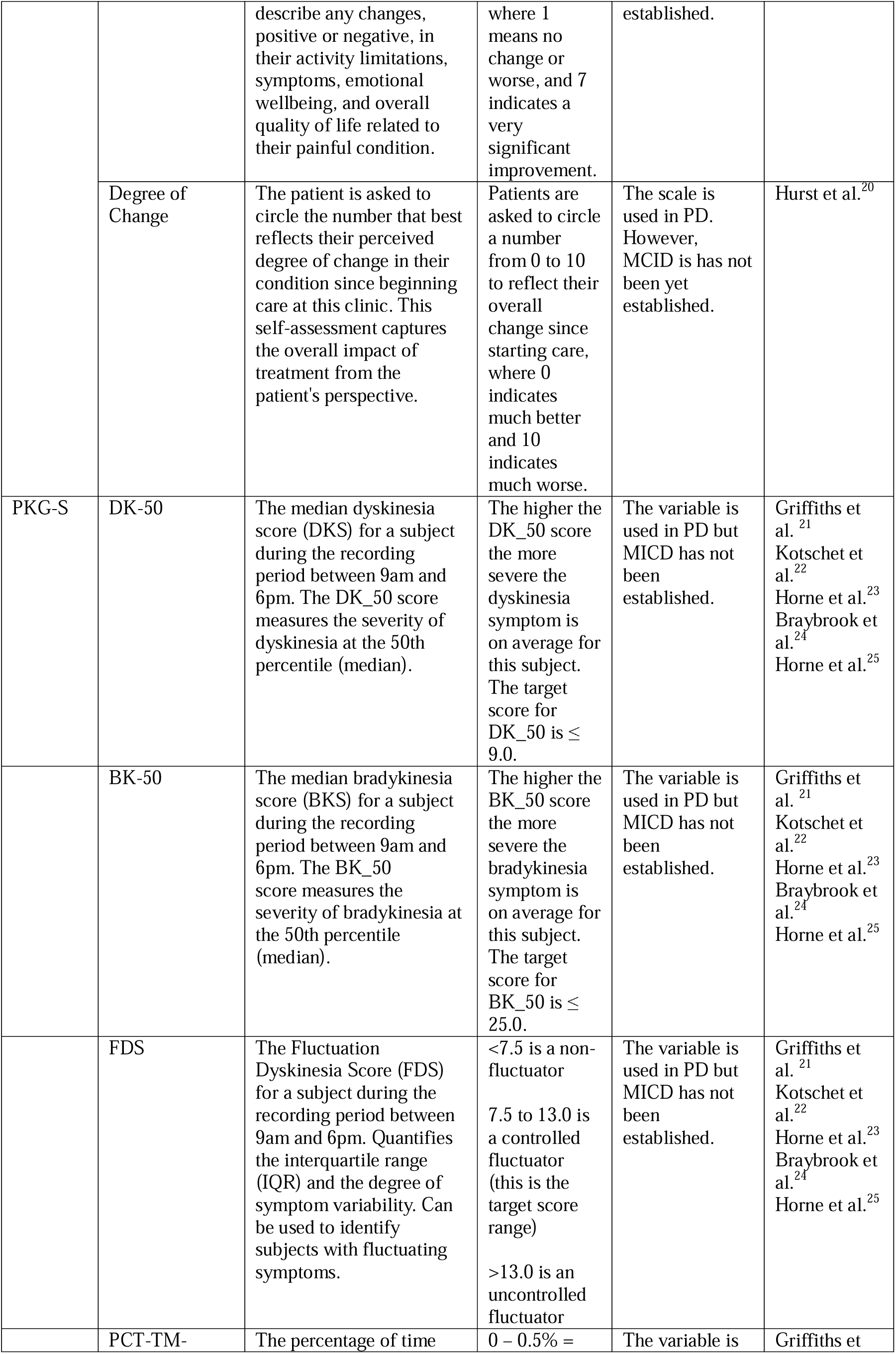
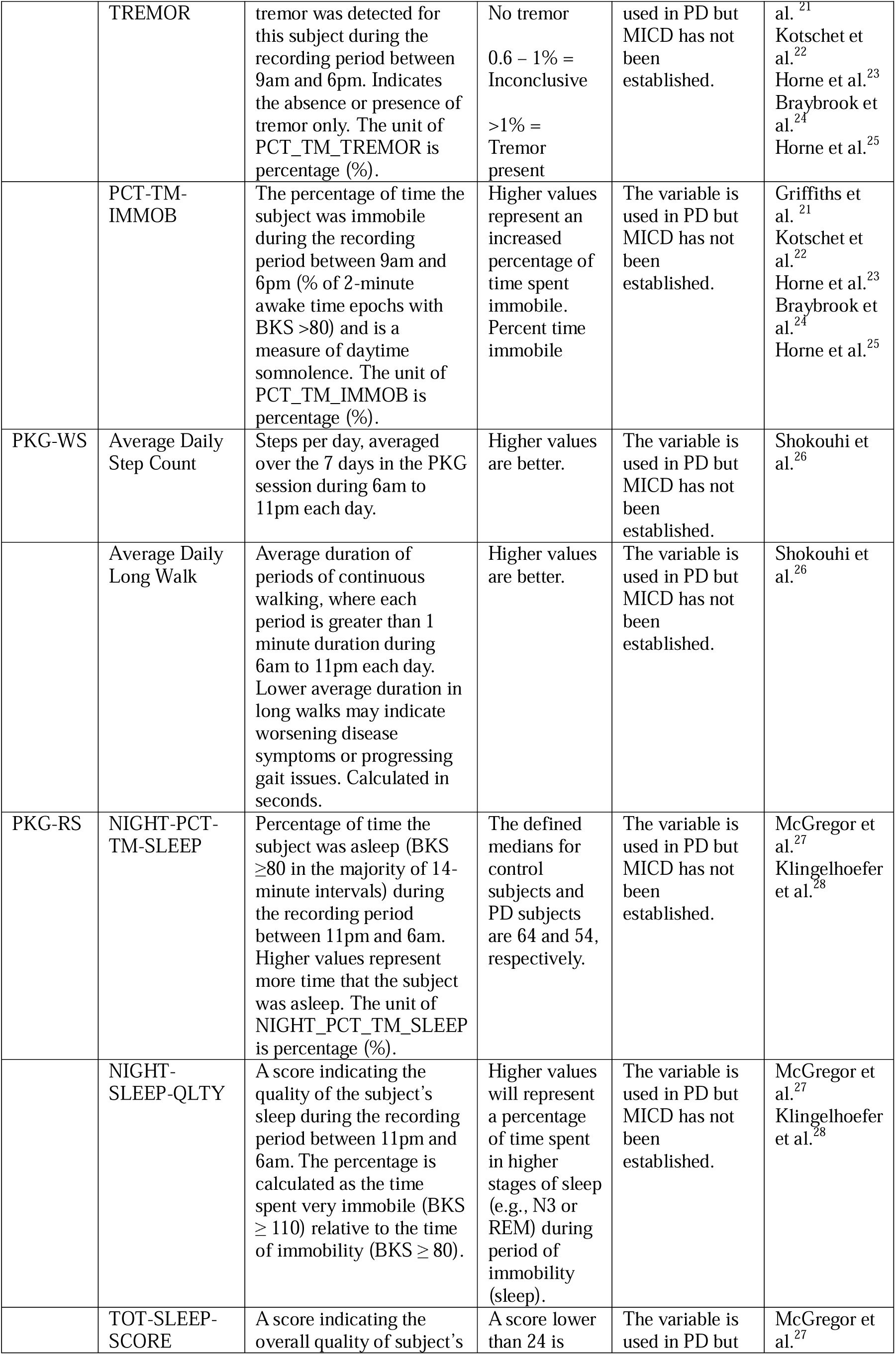
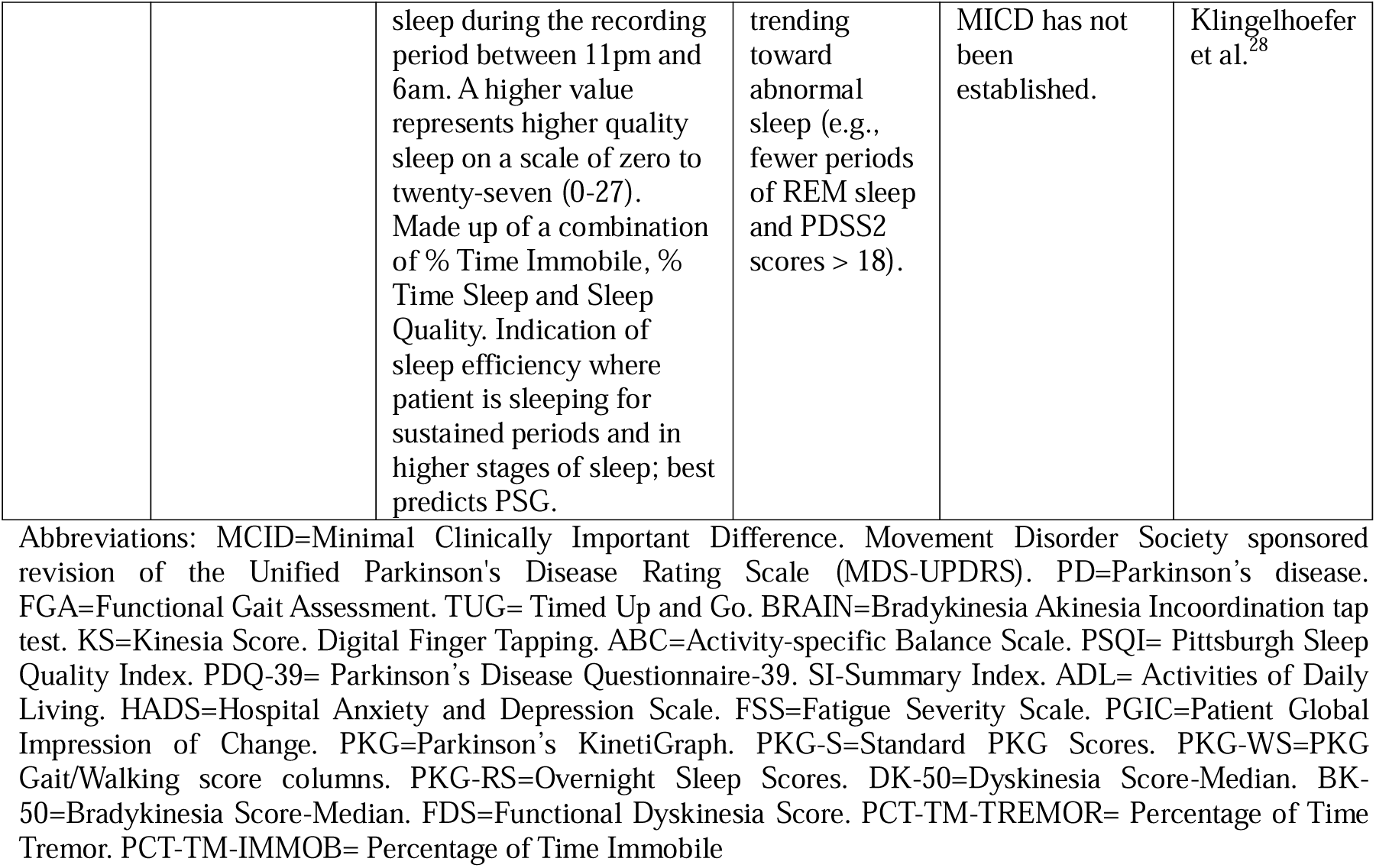

## Appendix 4: Between-group baseline scores for clinical effectiveness outcomes

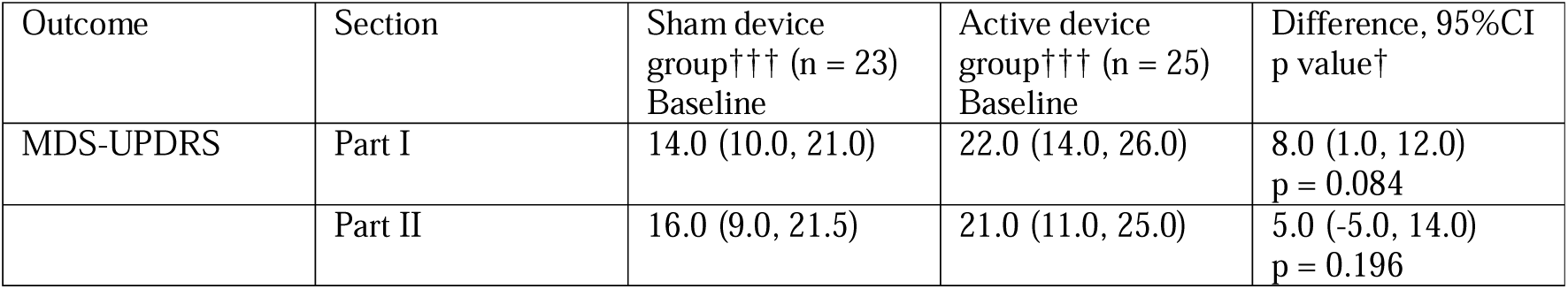
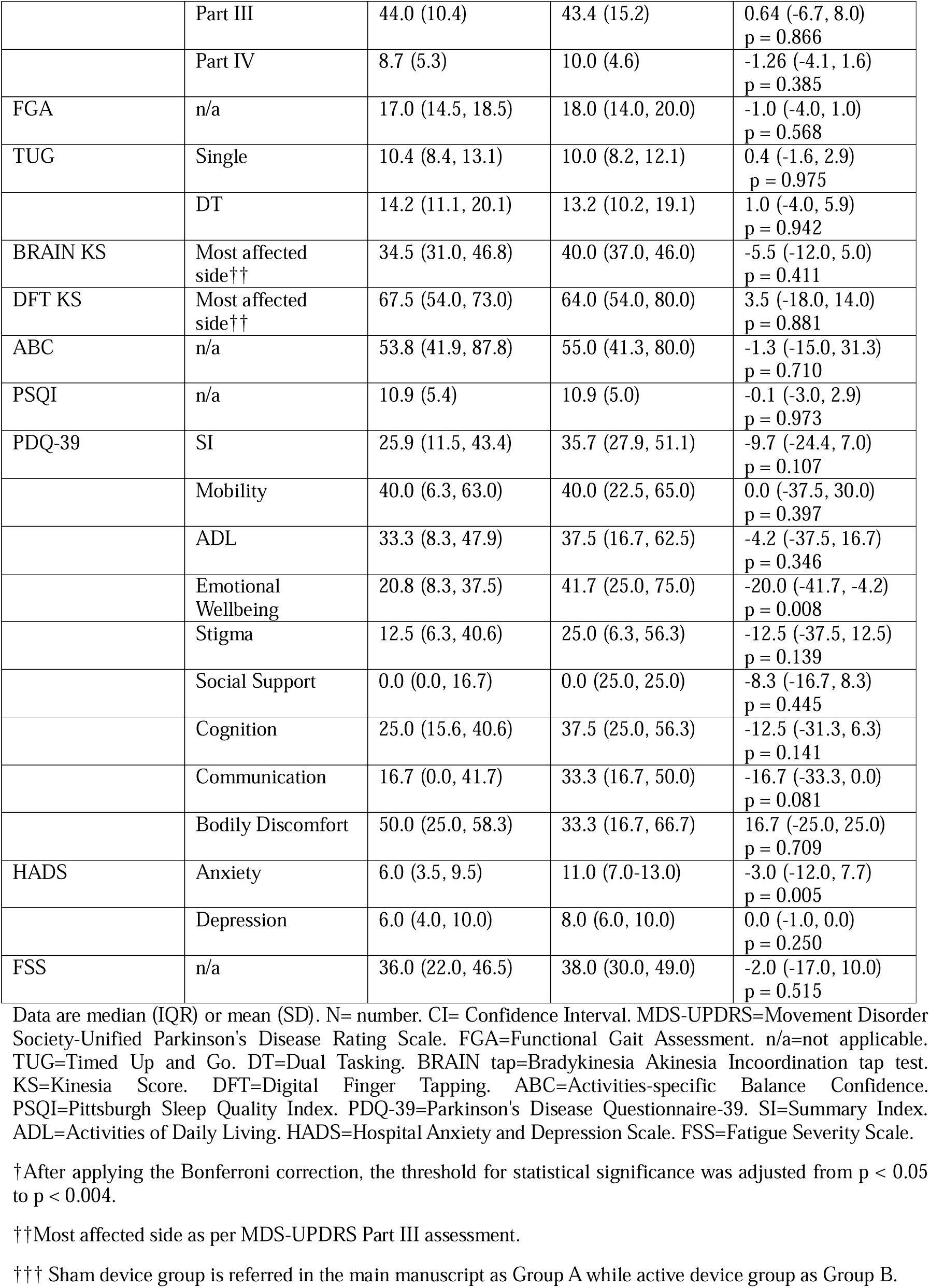

| Outcome | Section | Sham device<br>group††† (n = 23)<br>Baseline | Active device<br>group††† (n = 25)<br>Baseline | Difference, 95%CI<br>p value† |
| --- | --- | --- | --- | --- |
| MDS-UPDRS | Part I | 14.0 (10.0, 21.0) | 22.0 (14.0, 26.0) | 8.0 (1.0, 12.0)<br>p = 0.084 |
|  | Part II | 16.0 (9.0, 21.5) | 21.0 (11.0, 25.0) | 5.0 (-5.0, 14.0)<br>p = 0.196 |
|  | Part III | 44.0 (10.4) | 43.4 (15.2) | 0.64 (-6.7, 8.0)<br>p = 0.866 |
|  | Part IV | 8.7 (5.3) | 10.0 (4.6) | -1.26 (-4.1, 1.6)<br>p = 0.385 |
| FGA | n/a | 17.0 (14.5, 18.5) | 18.0 (14.0, 20.0) | -1.0 (-4.0, 1.0)<br>p = 0.568 |
| TUG | Single | 10.4 (8.4, 13.1) | 10.0 (8.2, 12.1) | 0.4 (-1.6, 2.9)<br>p = 0.975 |
|  | DT | 14.2 (11.1, 20.1) | 13.2 (10.2, 19.1) | 1.0 (-4.0, 5.9)<br>p = 0.942 |
| BRAIN KS | Most affected side†† | 34.5 (31.0, 46.8) | 40.0 (37.0, 46.0) | -5.5 (-12.0, 5.0)<br>p = 0.411 |
| DFT KS | Most affected side†† | 67.5 (54.0, 73.0) | 64.0 (54.0, 80.0) | 3.5 (-18.0, 14.0)<br>p = 0.881 |
| ABC | n/a | 53.8 (41.9, 87.8) | 55.0 (41.3, 80.0) | -1.3 (-15.0, 31.3)<br>p = 0.710 |
| PSQI | n/a | 10.9 (5.4) | 10.9 (5.0) | -0.1 (-3.0, 2.9)<br>p = 0.973 |
| PDQ-39 | SI | 25.9 (11.5, 43.4) | 35.7 (27.9, 51.1) | -9.7 (-24.4, 7.0)<br>p = 0.107 |
|  | Mobility | 40.0 (6.3, 63.0) | 40.0 (22.5, 65.0) | 0.0 (-37.5, 30.0)<br>p = 0.397 |
|  | ADL | 33.3 (8.3, 47.9) | 37.5 (16.7, 62.5) | -4.2 (-37.5, 16.7)<br>p = 0.346 |
|  | Emotional Wellbeing | 20.8 (8.3, 37.5) | 41.7 (25.0, 75.0) | -20.0 (-41.7, -4.2)<br>p = 0.008 |
|  | Stigma | 12.5 (6.3, 40.6) | 25.0 (6.3, 56.3) | -12.5 (-37.5, 12.5)<br>p = 0.139 |
|  | Social Support | 0.0 (0.0, 16.7) | 0.0 (25.0, 25.0) | -8.3 (-16.7, 8.3)<br>p = 0.445 |
|  | Cognition | 25.0 (15.6, 40.6) | 37.5 (25.0, 56.3) | -12.5 (-31.3, 6.3)<br>p = 0.141 |
|  | Communication | 16.7 (0.0, 41.7) | 33.3 (16.7, 50.0) | -16.7 (-33.3, 0.0)<br>p = 0.081 |
|  | Bodily Discomfort | 50.0 (25.0, 58.3) | 33.3 (16.7, 66.7) | 16.7 (-25.0, 25.0)<br>p = 0.709 |
| HADS | Anxiety | 6.0 (3.5, 9.5) | 11.0 (7.0-13.0) | -3.0 (-12.0, 7.7)<br>p = 0.005 |
|  | Depression | 6.0 (4.0, 10.0) | 8.0 (6.0, 10.0) | 0.0 (-1.0, 0.0)<br>p = 0.250 |
| FSS | n/a | 36.0 (22.0, 46.5) | 38.0 (30.0, 49.0) | -2.0 (-17.0, 10.0)<br>p = 0.515 |

Data are median (IQR) or mean (SD). N= number. CI= Confidence Interval. MDS-UPDRS=Movement Disorder Society-Unified Parkinson's Disease Rating Scale. FGA=Functional Gait Assessment. n/a=not applicable. TUG=Timed Up and Go. DT=Dual Tasking. BRAIN tap=Bradykinesia Akinesia Incoordination tap test. KS=Kinesia Score. DFT=Digital Finger Tapping. ABC=Activities-specific Balance Confidence. PSQI=Pittsburgh Sleep Quality Index. PDQ-39=Parkinson's Disease Questionnaire-39. SI=Summary Index. ADL=Activities of Daily Living. HADS=Hospital Anxiety and Depression Scale. FSS=Fatigue Severity Scale.
†After applying the Bonferroni correction, the threshold for statistical significance was adjusted from $p < 0.05$ to $p < 0.004$ .
††Most affected side as per MDS-UPDRS Part III assessment.
††† Sham device group is referred in the main manuscript as Group A while active device group as Group B.

## Appendix 5: Between-group baseline scores for Parkinson’s KinetiGraph outcomes

***Appendix 5: Between-group baseline scores for Parkinson's KinetiGraph outcomes.***
| Outcome | Section | Sham device<br>group†† (n = 23)<br>Baseline | Active device<br>group†† (n = 25)<br>Baseline | Difference,<br>95%CI<br>p value† |
| --- | --- | --- | --- | --- |
| PKG-S | DK-50 | 1.4<br>(0.4, 1.9) | 1.4<br>(0.9, 3.1) | 0.0<br>(-54.4, 9.4)<br>p = 0.300 |
|  | BK-50 | 29.2<br>(24.6, 36.4) | 28.1<br>(26.6, 32.6) | -1.1<br>(-19.0, 20.9)<br>p = 0.337 |
|  | FDS | 8.1<br>(6.4, 10.1) | 8.0<br>(6.4, 10.3) | -0.1<br>(-2.1, 2.4)<br>p = 0.856 |
|  | PCT-TM-TREMOR | 2.7<br>(0.9, 5.4) | 1.8<br>(1.1, 6.4) | -0.9<br>(-3.5, 3.3)<br>p = 0.957 |
|  | PCT-TM-IMMOB | 9.0<br>(5.2, 15.5) | 10.5<br>(5.9, 14.9) | 1.6<br>(-6.7, 7.0)<br>p = 0.957 |
| PKG-WS | Average Daily Step Count | 4127.0<br>(2706.0, 5828.8) | 5528.0<br>(3807.0, 8390.0) | 1401.0<br>(-887.5, 4128.0)<br>p = 0.321 |
|  | Average Daily Long Walk | 1057.0<br>(339.8, 1676.3) | 737.0<br>(416.0, 2026.0) | -320<br>(-750.5, 771.5)<br>p = 0.941 |
| PKG-RS | NIGHT-PCT-TM-SLEEP | 44.3<br>(35.5, 56.4) | 60.1<br>(46.8, 73.4) | 15.8<br>(1.9, 32.6)<br>p = 0.026 |
|  | NIGHT-SLEEP-QLTY | 59.0<br>(49.8, 75.4) | 73.6<br>(58.6, 84.1) | 14.7<br>(-0.8, 27.5)<br>p = 0.044 |
|  | TOT-SLEEP-SCORE | 1.5<br>(0.0, 6.3) | 7.0<br>(3.0, 21.0) | 5.5<br>(1.5, 18.0)<br>p = 0.026 |

Data are median (IQR) or mean (SD). N= number. CI= Confidence Interval. PKG=Parkinson's KinetiGraph. PKG-S=Standard PKG Scores. PKG-WS=PKG Gait/Walking score columns. PKG-RS=Overnight Sleep Scores. DK-50=Dyskinesia Score-Median. BK-50=Bradykinesia Score-Median. FDS=Functional Dyskinesia Score. PCT-TM-TREMOR= Percentage of Time Tremor. PCT-TM-IMMOB= Percentage of Time Immobile. PKG wrist watch was worn on most affected side.
†After applying the Bonferroni correction, the threshold for statistical significance was adjusted from $p < 0.05$ to $p < 0.004$ .
†† Sham device group is referred in the main manuscript as Group A while active device group as Group B.

## Appendix 6: Between group change in outcomes at follow up

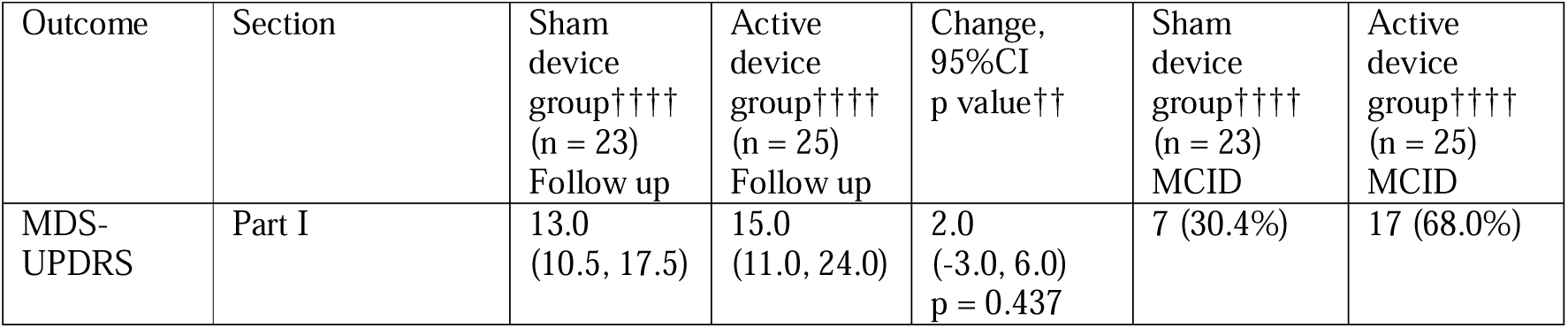
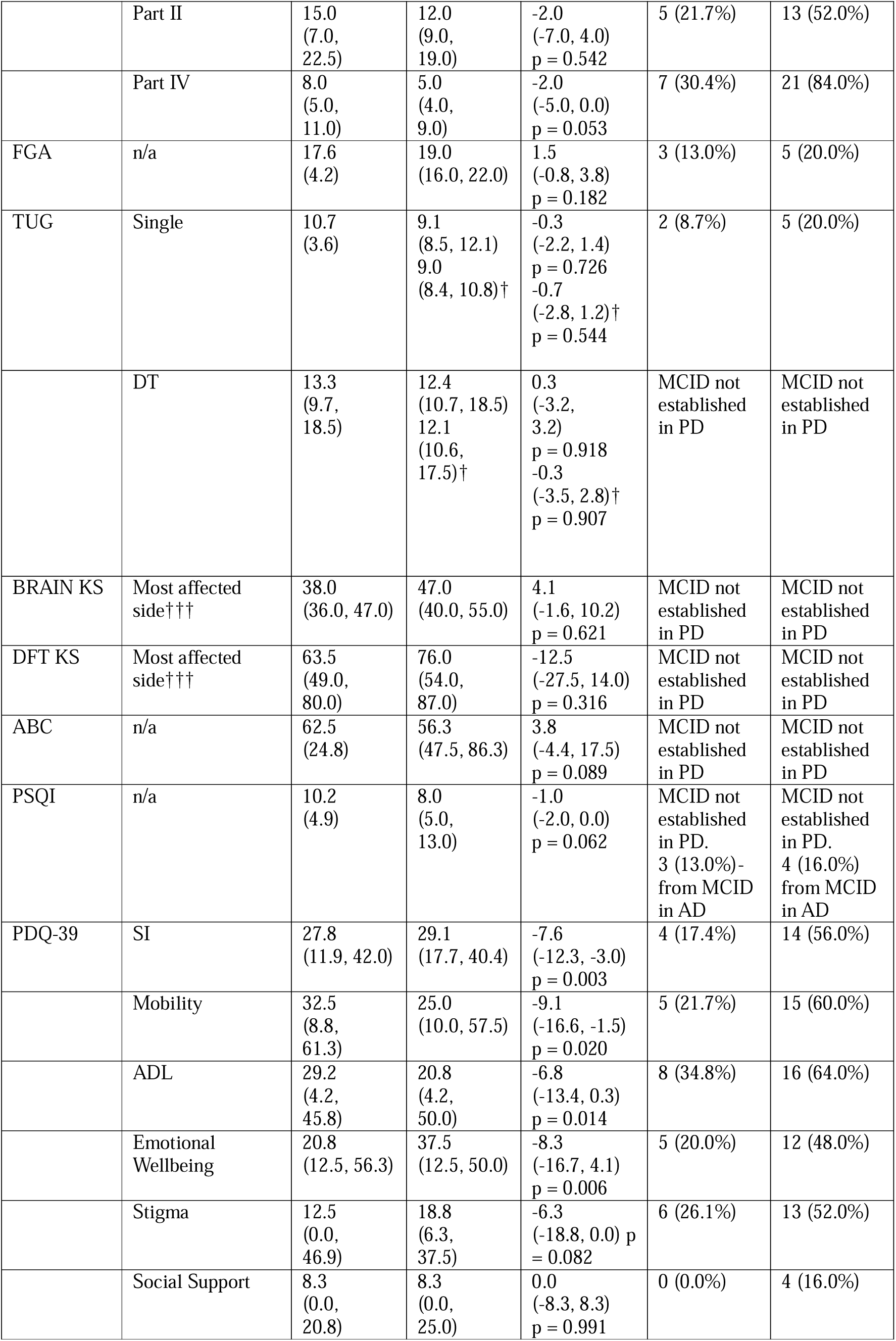
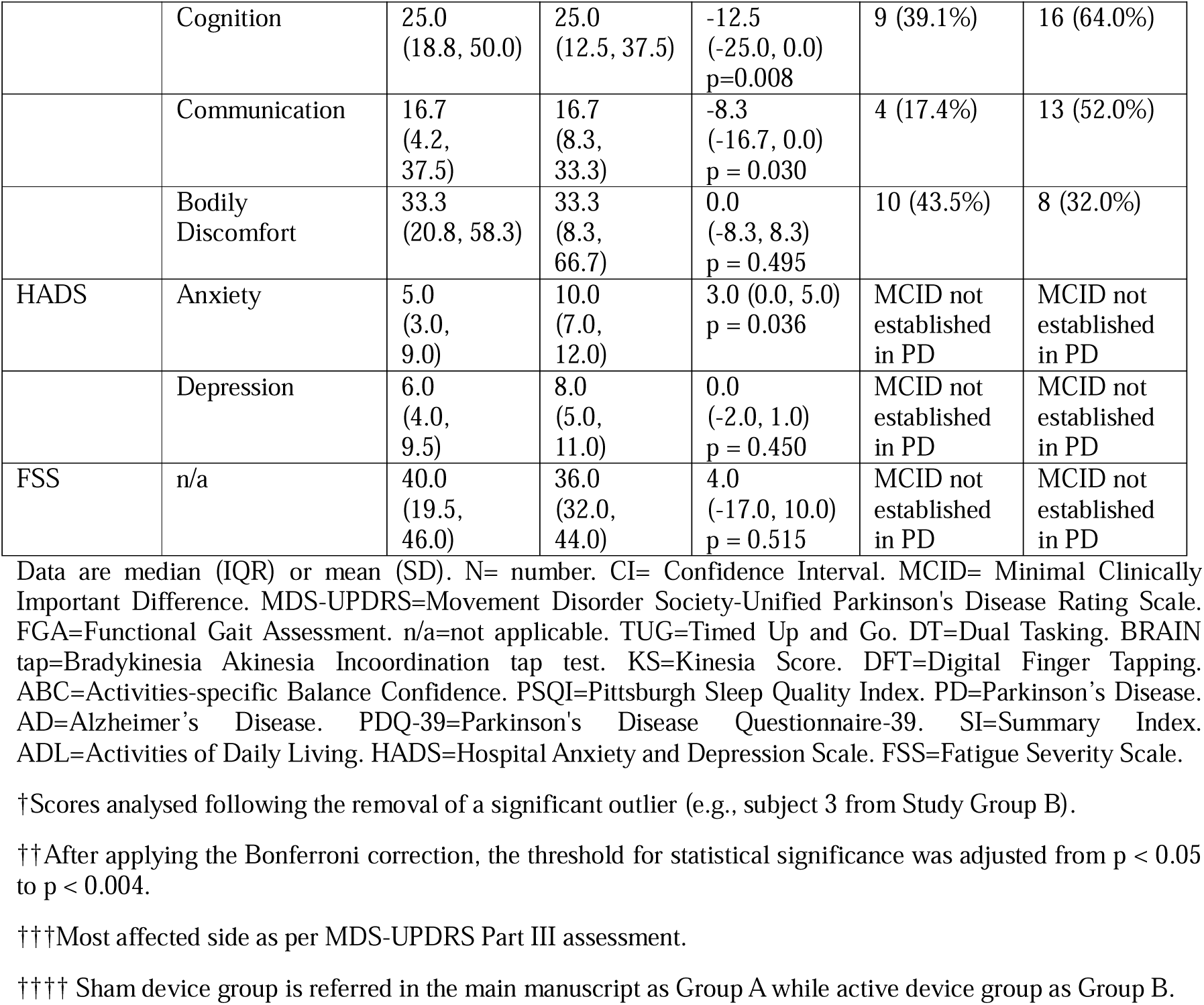

**Appendix 6: Between group change in outcomes at follow up.**
| Outcome | Section | Sham device group††††<br>(n = 23)<br>Follow up | Active device group††††<br>(n = 25)<br>Follow up | Change,<br>95%CI<br>p value†† | Sham device group††††<br>(n = 23)<br>MCID | Active device group††††<br>(n = 25)<br>MCID |
| --- | --- | --- | --- | --- | --- | --- |
| MDS-UPDRS | Part I | 13.0<br>(10.5, 17.5) | 15.0<br>(11.0, 24.0) | 2.0<br>(-3.0, 6.0)<br>p = 0.437 | 7 (30.4%) | 17 (68.0%) |
|  | Part II | 15.0<br>(7.0, 22.5) | 12.0<br>(9.0, 19.0) | -2.0<br>(-7.0, 4.0)<br>p = 0.542 | 5 (21.7%) | 13 (52.0%) |
|  | Part IV | 8.0<br>(5.0, 11.0) | 5.0<br>(4.0, 9.0) | -2.0<br>(-5.0, 0.0)<br>p = 0.053 | 7 (30.4%) | 21 (84.0%) |
| FGA | n/a | 17.6<br>(4.2) | 19.0<br>(16.0, 22.0) | 1.5<br>(-0.8, 3.8)<br>p = 0.182 | 3 (13.0%) | 5 (20.0%) |
| TUG | Single | 10.7<br>(3.6) | 9.1<br>(8.5, 12.1)<br>9.0<br>(8.4, 10.8)† | -0.3<br>(-2.2, 1.4)<br>p = 0.726<br>-0.7<br>(-2.8, 1.2)†<br>p = 0.544 | 2 (8.7%) | 5 (20.0%) |
|  | DT | 13.3<br>(9.7, 18.5) | 12.4<br>(10.7, 18.5)<br>12.1<br>(10.6, 17.5)† | 0.3<br>(-3.2, 3.2)<br>p = 0.918<br>-0.3<br>(-3.5, 2.8)†<br>p = 0.907 | MCID not established in PD | MCID not established in PD |
| BRAIN KS | Most affected side††† | 38.0<br>(36.0, 47.0) | 47.0<br>(40.0, 55.0) | 4.1<br>(-1.6, 10.2)<br>p = 0.621 | MCID not established in PD | MCID not established in PD |
| DFT KS | Most affected side††† | 63.5<br>(49.0, 80.0) | 76.0<br>(54.0, 87.0) | -12.5<br>(-27.5, 14.0)<br>p = 0.316 | MCID not established in PD | MCID not established in PD |
| ABC | n/a | 62.5<br>(24.8) | 56.3<br>(47.5, 86.3) | 3.8<br>(-4.4, 17.5)<br>p = 0.089 | MCID not established in PD | MCID not established in PD |
| PSQI | n/a | 10.2<br>(4.9) | 8.0<br>(5.0, 13.0) | -1.0<br>(-2.0, 0.0)<br>p = 0.062 | MCID not established in PD.<br>3 (13.0%)-<br>from MCID in AD | MCID not established in PD.<br>4 (16.0%)<br>from MCID in AD |
| PDQ-39 | SI | 27.8<br>(11.9, 42.0) | 29.1<br>(17.7, 40.4) | -7.6<br>(-12.3, -3.0)<br>p = 0.003 | 4 (17.4%) | 14 (56.0%) |
|  | Mobility | 32.5<br>(8.8, 61.3) | 25.0<br>(10.0, 57.5) | -9.1<br>(-16.6, -1.5)<br>p = 0.020 | 5 (21.7%) | 15 (60.0%) |
|  | ADL | 29.2<br>(4.2, 45.8) | 20.8<br>(4.2, 50.0) | -6.8<br>(-13.4, 0.3)<br>p = 0.014 | 8 (34.8%) | 16 (64.0%) |
|  | Emotional Wellbeing | 20.8<br>(12.5, 56.3) | 37.5<br>(12.5, 50.0) | -8.3<br>(-16.7, 4.1)<br>p = 0.006 | 5 (20.0%) | 12 (48.0%) |
|  | Stigma | 12.5<br>(0.0, 46.9) | 18.8<br>(6.3, 37.5) | -6.3<br>(-18.8, 0.0) p = 0.082 | 6 (26.1%) | 13 (52.0%) |
|  | Social Support | 8.3<br>(0.0, 20.8) | 8.3<br>(0.0, 25.0) | 0.0<br>(-8.3, 8.3)<br>p = 0.991 | 0 (0.0%) | 4 (16.0%) |
|  | Cognition | 25.0<br>(18.8, 50.0) | 25.0<br>(12.5, 37.5) | -12.5<br>(-25.0, 0.0)<br>p=0.008 | 9 (39.1%) | 16 (64.0%) |
|  | Communication | 16.7<br>(4.2, 37.5) | 16.7<br>(8.3, 33.3) | -8.3<br>(-16.7, 0.0)<br>p = 0.030 | 4 (17.4%) | 13 (52.0%) |
|  | Bodily Discomfort | 33.3<br>(20.8, 58.3) | 33.3<br>(8.3, 66.7) | 0.0<br>(-8.3, 8.3)<br>p = 0.495 | 10 (43.5%) | 8 (32.0%) |
| HADS | Anxiety | 5.0<br>(3.0, 9.0) | 10.0<br>(7.0, 12.0) | 3.0 (0.0, 5.0)<br>p = 0.036 | MCID not established in PD | MCID not established in PD |
|  | Depression | 6.0<br>(4.0, 9.5) | 8.0<br>(5.0, 11.0) | 0.0<br>(-2.0, 1.0)<br>p = 0.450 | MCID not established in PD | MCID not established in PD |
| FSS | n/a | 40.0<br>(19.5, 46.0) | 36.0<br>(32.0, 44.0) | 4.0<br>(-17.0, 10.0)<br>p = 0.515 | MCID not established in PD | MCID not established in PD |

Data are median (IQR) or mean (SD). N= number. CI= Confidence Interval. MCID= Minimal Clinically Important Difference. MDS-UPDRS=Movement Disorder Society-Unified Parkinson's Disease Rating Scale. FGA=Functional Gait Assessment. n/a=not applicable. TUG=Timed Up and Go. DT=Dual Tasking. BRAIN tap=Bradykinesia Akinesia Incoordination tap test. KS=Kinesia Score. DFT=Digital Finger Tapping. ABC=Activities-specific Balance Confidence. PSQI=Pittsburgh Sleep Quality Index. PD=Parkinson's Disease. AD=Alzheimer's Disease. PDQ-39=Parkinson's Disease Questionnaire-39. SI=Summary Index. ADL=Activities of Daily Living. HADS=Hospital Anxiety and Depression Scale. FSS=Fatigue Severity Scale.
†Scores analysed following the removal of a significant outlier (e.g., subject 3 from Study Group B).
††After applying the Bonferroni correction, the threshold for statistical significance was adjusted from $p < 0.05$ to $p < 0.004$ .
†††Most affected side as per MDS-UPDRS Part III assessment.
†††† Sham device group is referred in the main manuscript as Group A while active device group as Group B.

## Appendix 7: Within group change in outcomes at follow up

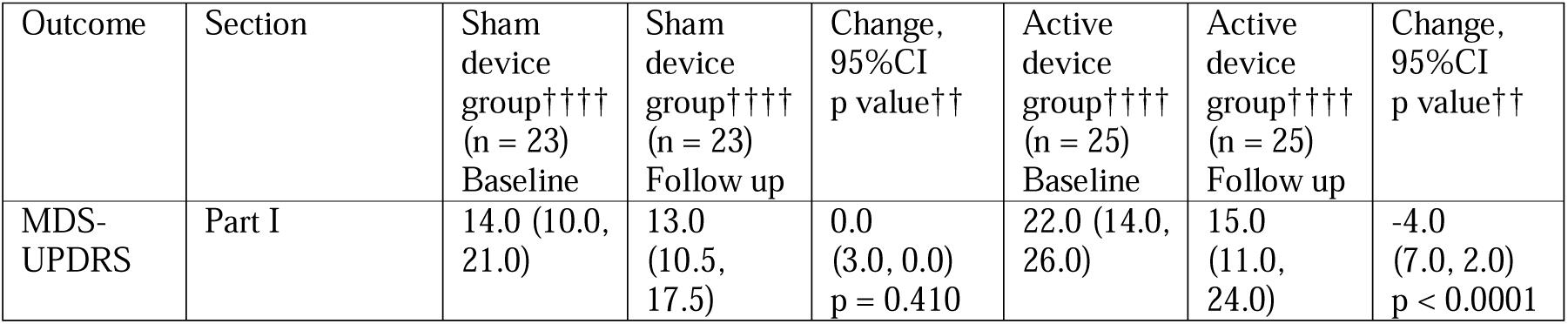
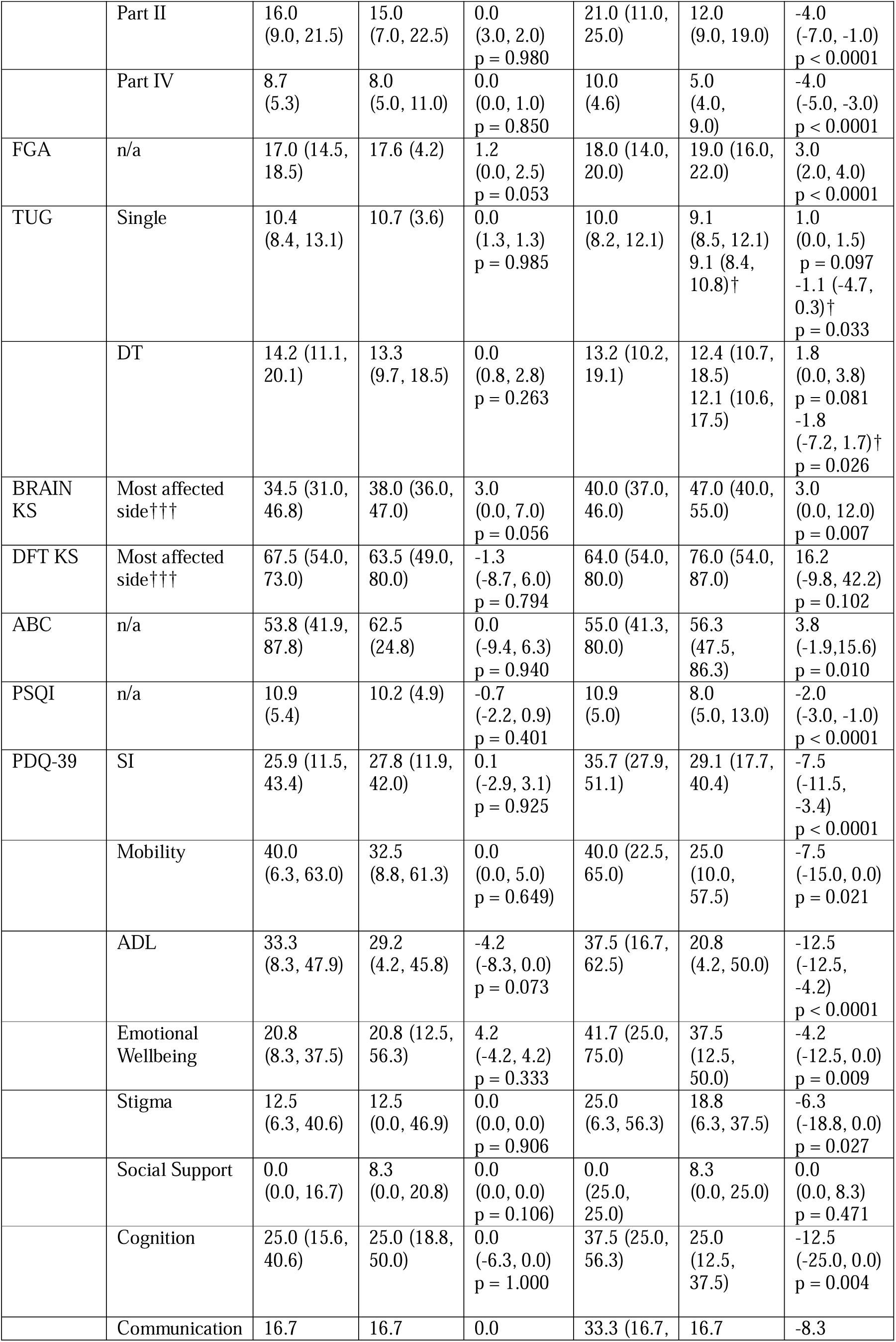
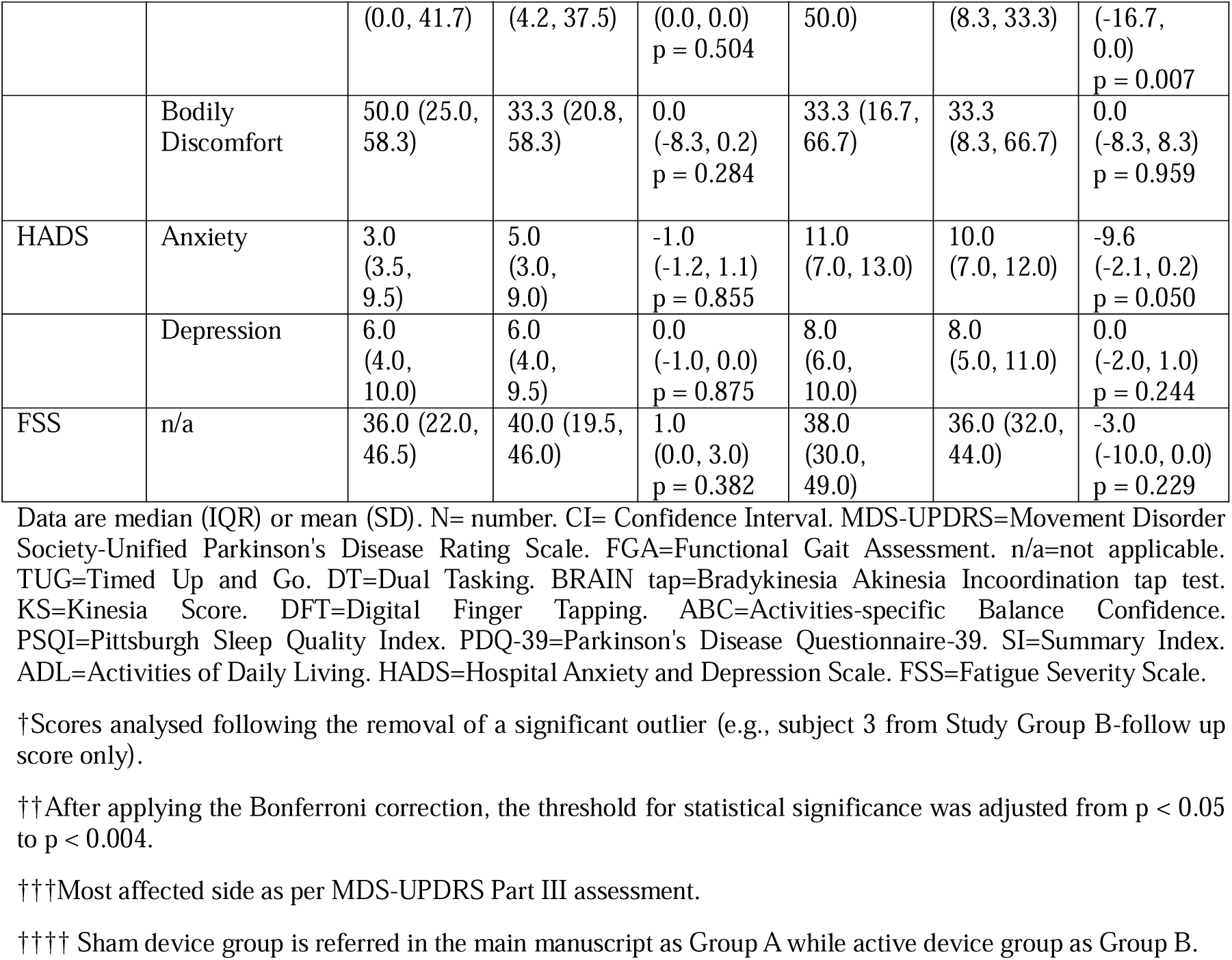

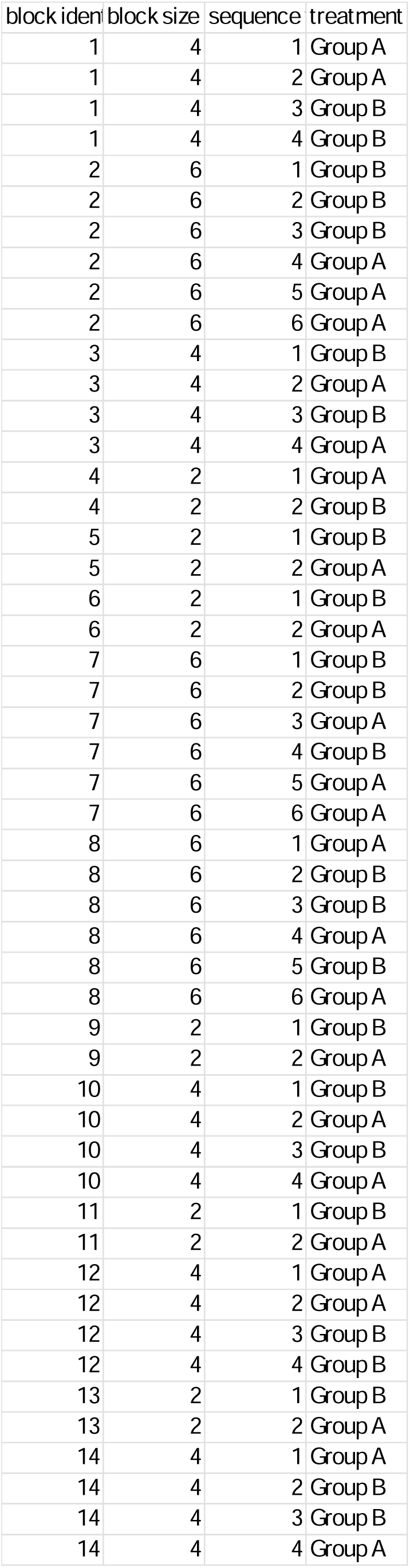

## References

1. GBD 2017 US Neurological Disorders Collaborators; Feigin VL, Vos T, Alahdab F, Amit AML, Bärnighausen TW, Beghi E, Beheshti M, Chavan PP, Criqui MH, Desai R, Dhamminda Dharmaratne S, Dorsey ER, Wilder Eagan A, Elgendy IY, Filip I, Giampaoli S, Giussani G, Hafezi-Nejad N, Hole MK, Ikeda T, Owens Johnson C, Kalani R, Khatab K, Khubchandani J, Kim D, Koroshetz WJ, Krishnamoorthy V, Krishnamurthi RV, Liu X, Lo WD, Logroscino G, Mensah GA, Miller TR, Mohammed S, Mokdad AH, Moradi-Lakeh M, Morrison SD, Shivamurthy VKN, Naghavi M, Nichols E, Norrving B, Odell CM, Pupillo E, Radfar A, Roth GA, Shafieesabet A, Sheikh A, Sheikhbahaei S, Shin JI, Singh JA, Steiner TJ, Stovner LJ, Wallin MT, Weiss J, Wu C, Zunt JR, Adelson JD, Murray CJL. Burden of Neurological Disorders Across the US From 1990-2017: A Global Burden of Disease Study. JAMA Neurol. 2021 Feb 1;78(2):165–176. doi: 10.1001/jamaneurol.2020.4152. PMID: 33136137; PMCID: PMC7607495.

2. Bloem BR, Okun MS, Klein C. Parkinson’s disease. Lancet. 2021 Jun 12;397(10291):2284-2303. doi: 10.1016/S0140-6736(21)00218-X. Epub 2021 Apr 10. PMID: 33848468.

3. Fahn S, Oakes D, Shoulson I, Kieburtz K, Rudolph A, Lang A, Olanow CW, Tanner C, Marek K; Parkinson Study Group. Levodopa and the progression of Parkinson’s disease. N Engl J Med. 2004 Dec 9;351(24):2498–508. doi: 10.1056/NEJMoa033447. PMID: 15590952.

4. Masood N, Jimenez-Shahed J. Effective Management of “OFF” Episodes in Parkinson’s Disease: Emerging Treatment Strategies and Unmet Clinical Needs. Neuropsychiatr Dis Treat. 2023 Jan 25;19:247–266. doi: 10.2147/NDT.S273121. PMID: 36721795; PMCID: PMC9884436.

5. Dorsey ER, Bloem BR. The Parkinson Pandemic-A Call to Action. JAMA Neurol. 2018 Jan 1;75(1):9–10. doi: 10.1001/jamaneurol.2017.3299. PMID: 29131880.

6. Olanow CW, Obeso JA, Stocchi F. Continuous dopamine-receptor treatment of Parkinson’s disease: scientific rationale and clinical implications. Lancet Neurol. 2006 Aug;5(8):677–87. doi: 10.1016/S1474-4422(06)70521-X. PMID: 16857573.

7. Deuschl G, Schade-Brittinger C, Krack P, Volkmann J, Schäfer H, Bötzel K, Daniels C, Deutschländer A, Dillmann U, Eisner W, Gruber D, Hamel W, Herzog J, Hilker R, Klebe S, Kloss M, Koy J, Krause M, Kupsch A, Lorenz D, Lorenzl S, Mehdorn HM, Moringlane JR, Oertel W, Pinsker MO, Reichmann H, Reuss A, Schneider GH, Schnitzler A, Steude U, Sturm V, Timmermann L, Tronnier V, Trottenberg T, Wojtecki L, Wolf E, Poewe W, Voges J; German Parkinson Study Group, Neurostimulation Section. A randomized trial of deep-brain stimulation for Parkinson’s disease. N Engl J Med. 2006 Aug 31;355(9):896–908. doi: 10.1056/NEJMoa060281. Erratum in: N Engl J Med. 2006 Sep 21;355(12):1289. PMID: 16943402.

8. Rees RN, Noyce AJ, Schrag AE. Identification of Prodromal Parkinson Disease: We May Be Able to But Should We? Neurology. 2024 Jun 11;102(11):e209394. doi: 10.1212/WNL.0000000000209394. Epub 2024 May 17. PMID: 38759130; PMCID: PMC11175649.

9. Azoidou V, Rowsell K, Camboe E, Dey KC, Zirra A, Quah C, Boyle T, Gallagher D, Noyce AJ, Simonet C. A pilot interventional study on feasibility and effectiveness of the CUE1 device in Parkinson’s disease. Parkinsonism Relat Disord. 2025 Apr;133:107349. doi: 10.1016/j.parkreldis.2025.107349. Epub 2025 Feb 22. PMID: 40015168.

10. Azoidou V, Bhadra E, Rowsell K, Camboe E, Dey K, Zirra A, Quah C, Budu C, Boyle T, Gallagher D, Noyce A, Simonet C. Non-invasive device to alleviate symptoms in people living with Parkinson’s: study protocol for a multicentre phase II double-blind randomised controlled trial. BMJ Open. 2025 Apr 27;15(4):e096051. doi: 10.1136/bmjopen-2024-096051. PMID: 40288787; PMCID: PMC12035462.

11. Azoidou V, Noyce AJ, Simonet C. The effect of tactile cueing on dual task performance in Parkinson’s disease. A systematic review and meta-analysis. Clin Park Relat Disord. 2024 Nov 17;11:100284. doi: 10.1016/j.prdoa.2024.100284. PMID: 39640985; PMCID: PMC11617393.

12. Cosentino C, Putzolu M, Mezzarobba S, Cecchella M, Innocenti T, Bonassi G, Botta A, Lagravinese G, Avanzino L, Pelosin E. One cue does not fit all: A systematic review with meta-analysis of the effectiveness of cueing on freezing of gait in Parkinson’s disease. Neurosci Biobehav Rev. 2023 Jul;150:105189. doi: 10.1016/j.neubiorev.2023.105189. Epub 2023 Apr 20. PMID: 37086934.

13. Klaver EC, van Vugt JPP, Bloem BR, van Wezel RJA, Nonnekes J, Tjepkema-Cloostermans MC. Good vibrations: tactile cueing for freezing of gait in Parkinson’s disease. J Neurol. 2023 Jul;270(7):3424–3432. doi: 10.1007/s00415-023-11663-9. Epub 2023 Mar 21. PMID: 36944760; PMCID: PMC10267272.

14. Volpe D, Giantin MG, Fasano A. A wearable proprioceptive stabilizer (Equistasi®) for rehabilitation of postural instability in Parkinson’s disease: a phase II randomized double-blind, double-dummy, controlled study. PLoS One. 2014 Nov 17;9(11):e112065. doi: 10.1371/journal.pone.0112065. PMID: 25401967; PMCID: PMC4234681.

15. Serio F, Minosa C, De Luca M, Conte P, Albani G, Peppe A. Focal Vibration Training (Equistasi^®^) to Improve Posture Stability. A Retrospective Study in Parkinson’s Disease. Sensors (Basel). 2019 May 7;19(9):2101. doi: 10.3390/s19092101. PMID: 31067663; PMCID: PMC6539920.

16. Tosserams A, Weerdesteyn V, Bal T, Bloem BR, Solis-Escalante T, Nonnekes J. Cortical Correlates of Gait Compensation Strategies in Parkinson Disease. Ann Neurol. 2022 Mar;91(3):329–341. doi: 10.1002/ana.26306. Epub 2022 Feb 8. PMID: 35067999; PMCID: PMC9306676.

17. National Institute for Health and Care Excellence (NICE). Evidence Standards Framework for Digital Health Technologies. 2023.

18. NHS England. Digital Health Evidence Generation Guide. 2023.

19. Rowling Clinic Public Engagement Report. Edinburgh: Anne Rowling Regenerative Neurology Clinic; 2023.

20. Postuma RB, Berg D, Stern M, Poewe W, Olanow CW, Oertel W, Obeso J, Marek K, Litvan I, Lang AE, Halliday G, Goetz CG, Gasser T, Dubois B, Chan P, Bloem BR, Adler CH, Deuschl G. MDS clinical diagnostic criteria for Parkinson’s disease. Mov Disord. 2015 Oct;30(12):1591–601. doi: 10.1002/mds.26424. PMID: 26474316.

21. Hoehn MM, Yahr MD. Parkinsonism: onset, progression and mortality. Neurology. 1967 May;17(5):427-42. doi: 10.1212/wnl.17.5.427. PMID: 6067254.

22. Hoops S, Nazem S, Siderowf AD, Duda JE, Xie SX, Stern MB, Weintraub D. Validity of the MoCA and MMSE in the detection of MCI and dementia in Parkinson disease. Neurology. 2009 Nov 24;73(21):1738–45. doi: 10.1212/WNL.0b013e3181c34b47. PMID: 19933974; PMCID: PMC2788810.

23. Office for National Statistics. Ethnic group classifications: Census 2021. Newport (UK): ONS; 2021 [cited 2025 Jun 9]. Available from: https://www.ons.gov.uk/peoplepopulationandcommunity/culturalidentity/ethnicity/bulletins/ethnicgroupenglandandwales/census2021

24. Goetz CG, Tilley BC, Shaftman SR, Stebbins GT, Fahn S, Martinez-Martin P, Poewe W, Sampaio C, Stern MB, Dodel R, Dubois B, Holloway R, Jankovic J, Kulisevsky J, Lang AE, Lees A, Leurgans S, LeWitt PA, Nyenhuis D, Olanow CW, Rascol O, Schrag A, Teresi JA, van Hilten JJ, LaPelle N; Movement Disorder Society UPDRS Revision Task Force. Movement Disorder Society-sponsored revision of the Unified Parkinson’s Disease Rating Scale (MDS-UPDRS): scale presentation and clinimetric testing results. Mov Disord. 2008 Nov 15;23(15):2129–70. doi: 10.1002/mds.22340. PMID: 19025984.

25. Petersen C, Steffen T, Paly E, Dvorak L, Nelson R. Reliability and Minimal Detectable Change for Sit-to-Stand Tests and the Functional Gait Assessment for Individuals With Parkinson Disease. J Geriatr Phys Ther. 2017 Oct/Dec;40(4):223–226. doi: 10.1519/JPT.0000000000000102. PMID: 27805924.

26. Podsiadlo D, Richardson S. The timed “Up & Go”: a test of basic functional mobility for frail elderly persons. J Am Geriatr Soc. 1991 Feb;39(2):142–8. doi: 10.1111/j.1532-5415.1991.tb01616.x. PMID: 1991946.

27. Noyce AJ, Nagy A, Acharya S, Hadavi S, Bestwick JP, Fearnley J, Lees AJ, Giovannoni G. Bradykinesia-akinesia incoordination test: validating an online keyboard test of upper limb function. PLoS One. 2014 Apr 29;9(4):e96260. doi: 10.1371/journal.pone.0096260. Erratum in: PLoS One. 2014;9(8):e105488. PMID: 24781810; PMCID: PMC4004565.

28. Akram N, Li H, Ben-Joseph A, Budu C, Gallagher DA, Bestwick JP, Schrag A, Noyce AJ, Simonet C. Developing and assessing a new web-based tapping test for measuring distal movement in Parkinson’s disease: a Distal Finger Tapping test. Sci Rep. 2022 Jan 10;12(1):386. doi: 10.1038/s41598-021-03563-7. PMID: 35013372; PMCID: PMC8748736.

29. Odin P, Ray Chaudhuri K, Slevin JT, Volkmann J, Dietrichs E, Martinez-Martin P, Krauss JK, Henriksen T, Katzenschlager R, Antonini A, Rascol O, Poewe W; National Steering Committees. Collective physician perspectives on non-oral medication approaches for the management of clinically relevant unresolved issues in Parkinson’s disease: Consensus from an international survey and discussion program. Parkinsonism Relat Disord. 2015 Oct;21(10):1133–44. doi: 10.1016/j.parkreldis.2015.07.020. Epub 2015 Jul 23. PMID: 26233582.

30. Mak MK, Pang MY. Fear of falling is independently associated with recurrent falls in patients with Parkinson’s disease: a 1-year prospective study. J Neurol. 2009 Oct;256(10):1689–95. doi: 10.1007/s00415-009-5184-5. Epub 2009 May 28. PMID: 19479166.

31. Buysse DJ, Reynolds CF 3rd, Monk TH, Berman SR, Kupfer DJ. The Pittsburgh Sleep Quality Index: a new instrument for psychiatric practice and research. Psychiatry Res. 1989 May;28(2):193–213. doi: 10.1016/0165-1781(89)90047-4. PMID: 2748771.

32. Jenkinson C, Fitzpatrick R, Peto V, Greenhall R, Hyman N. The Parkinson’s Disease Questionnaire (PDQ-39): development and validation of a Parkinson’s disease summary index score. Age Ageing. 1997 Sep;26(5):353–7. doi: 10.1093/ageing/26.5.353. PMID: 9351479.

33. Roberts SB, Bonnici DM, Mackinnon AJ, Worcester MC. Psychometric evaluation of the Hospital Anxiety and Depression Scale (HADS) among female cardiac patients. Br J Health Psychol. 2001 Nov;6(Part 4):373–383. doi: 10.1348/135910701169278. PMID: 12614511.

34. Hagell P, Höglund A, Reimer J, Eriksson B, Knutsson I, Widner H, Cella D. Measuring fatigue in Parkinson’s disease: a psychometric study of two brief generic fatigue questionnaires. J Pain Symptom Manage. 2006 Nov;32(5):420–32. doi: 10.1016/j.jpainsymman.2006.05.021. PMID: 17085268.

35. Hurst H, Bolton J. Assessing the clinical significance of change scores recorded on subjective outcome measures. J Manipulative Physiol Ther. 2004 Jan;27(1):26–35. doi: 10.1016/j.jmpt.2003.11.003. PMID: 14739871.

36. Copay AG, Subach BR, Glassman SD, Polly DW Jr, Schuler TC. Understanding the minimum clinically important difference: a review of concepts and methods. Spine J. 2007 Sep-Oct;7(5):541–6. doi: 10.1016/j.spinee.2007.01.008. Epub 2007 Apr 2. PMID: 17448732.

37. Sim J, Lewis M. The size of a pilot study for a clinical trial should be calculated in relation to considerations of precision and efficiency. J Clin Epidemiol. 2012 Mar;65(3):301–8. doi: 10.1016/j.jclinepi.2011.07.011. Epub 2011 Dec 9. PMID: 22169081.

38. Taichman DB, Sahni P, Pinborg A, Peiperl L, Laine C, James A, Hong ST, Haileamlak A, Gollogly L, Godlee F, Frizelle FA, Florenzano F, Drazen JM, Bauchner H, Baethge C, Backus J. Data Sharing Statements for Clinical Trials: A Requirement of the International Committee of Medical Journal Editors. Ann Intern Med. 2017 Jul 4;167(1):63–65. doi: 10.7326/M17-1028. Epub 2017 Jun 6. PMID: 28586790.

39. Horváth K, Aschermann Z, Ács P, Deli G, Janszky J, Komoly S, Balázs É, Takács K, Karádi K, Kovács N. Minimal clinically important difference on the Motor Examination part of MDS-UPDRS. Parkinsonism Relat Disord. 2015 Dec;21(12):1421–6. doi: 10.1016/j.parkreldis.2015.10.006. Epub 2015 Oct 22. PMID: 26578041.

40. Sánchez-Ferro Á, Matarazzo M, Martínez-Martín P, Martínez-Ávila JC, Gómez de la Cámara A, Giancardo L, Arroyo Gallego T, Montero P, Puertas-Martín V, Obeso I, Butterworth I, Mendoza CS, Catalán MJ, Molina JA, Bermejo-Pareja F, Martínez-Castrillo JC, López-Manzanares L, Alonso-Cánovas A, Herreros Rodríguez J, Gray M. Minimal Clinically Important Difference for UPDRS-III in Daily Practice. Mov Disord Clin Pract. 2018 Jun 26;5(4):448–450. doi: 10.1002/mdc3.12632. PMID: 30838303; PMCID: PMC6336372.

41. Fasano A, Canning CG, Hausdorff JM, Lord S, Rochester L. Falls in Parkinson’s disease: A complex and evolving picture. Mov Disord. 2017 Nov;32(11):1524–1536. doi: 10.1002/mds.27195. Epub 2017 Oct 25. PMID: 29067726.

42. Tang X, Yang J, Zhu Y, Gong H, Sun H, Chen F, Guan Q, Yu L, Wang W, Zhang Z, Li L, Ma G, Wang X. High PSQI score is associated with the development of dyskinesia in Parkinson’s disease. NPJ Parkinsons Dis. 2022 Sep 29;8(1):124. doi: 10.1038/s41531-022-00391-y. PMID: 36175559; PMCID: PMC9522669.

43. Simonet C, Pérez-Carbonell L, Galmés-Ordinas MA, Huxford BFR, Chohan H, Gill A, Leschziner G, Lees AJ, Schrag A, Noyce AJ. The Motor Dysfunction Seen in Isolated REM Sleep Behavior Disorder. Mov Disord. 2024 Jun;39(6):1054–1059. doi: 10.1002/mds.29779. Epub 2024 Mar 12. PMID: 38470080.

44. National Institute for Health and Care Excellence (NICE). Medical Technologies Evaluation Programme. London: NICE; [cited 2025 Jun 9]. Available from: https://www.nice.org.uk/about/what-we-do/our-programmes/nice-guidance/nice-medical-technologies-evaluation-programme

45. Rogers G, Davies D, Pink J, Cooper P. Parkinson’s disease: summary of updated NICE guidance. BMJ. 2017 Jul 27;358:j1951. doi: 10.1136/bmj.j1951. Erratum in: BMJ. 2019 Feb 28;364:l961. doi: 10.1136/bmj.l961. PMID: 28751362.

46. Schootemeijer S, van der Kolk NM, Ellis T, Mirelman A, Nieuwboer A, Nieuwhof F, Schwarzschild MA, de Vries NM, Bloem BR. Barriers and Motivators to Engage in Exercise for Persons with Parkinson’s Disease. J Parkinsons Dis. 2020;10(4):1293–1299. doi: 10.3233/JPD-202247. PMID: 32925106; PMCID: PMC7739964.

## Reference list

1. Goetz CG, Tilley BC, Shaftman SR, Stebbins GT, Fahn S, Martinez-Martin P, Poewe W, Sampaio C, Stern MB, Dodel R, Dubois B, Holloway R, Jankovic J, Kulisevsky J, Lang AE, Lees A, Leurgans S, LeWitt PA, Nyenhuis D, Olanow CW, Rascol O, Schrag A, Teresi JA, van Hilten JJ, LaPelle N; Movement Disorder Society UPDRS Revision Task Force. Movement Disorder Society-sponsored revision of the Unified Parkinson’s Disease Rating Scale (MDS-UPDRS): scale presentation and clinimetric testing results. Mov Disord. 2008 Nov 15;23(15):2129–70. doi: 10.1002/mds.22340. PMID: 19025984.

2. Horváth K, Aschermann Z, Kovács M, Makkos A, Harmat M, Janszky J, Komoly S, Karádi K, Kovács N. Minimal clinically important differences for the experiences of daily living parts of movement disorder society-sponsored unified Parkinson’s disease rating scale. Mov Disord. 2017 May;32(5):789–793. doi: 10.1002/mds.26960. Epub 2017 Feb 20. PMID: 28218413.

3. Horváth K, Aschermann Z, Ács P, Deli G, Janszky J, Komoly S, Balázs É, Takács K, Karádi K, Kovács N. Minimal clinically important difference on the Motor Examination part of MDS-UPDRS. Parkinsonism Relat Disord. 2015 Dec;21(12):1421–6. doi: 10.1016/j.parkreldis.2015.10.006. Epub 2015 Oct 22. PMID: 26578041.

4. Makkos A, Kovács M, Pintér D, Janszky J, Kovács N. Minimal clinically important difference for the historic parts of the Unified Dyskinesia Rating Scale. Parkinsonism Relat Disord. 2019 Jan;58:79–82. doi: 10.1016/j.parkreldis.2018.08.018. Epub 2018 Aug 26. PMID: 30174275.

5. Wrisley DM, Marchetti GF, Kuharsky DK, Whitney SL. Reliability, internal consistency, and validity of data obtained with the functional gait assessment. Phys Ther. 2004 Oct;84(10):906–18. PMID: 15449976.

6. Petersen C, Steffen T, Paly E, Dvorak L, Nelson R. Reliability and Minimal Detectable Change for Sit-to-Stand Tests and the Functional Gait Assessment for Individuals With Parkinson Disease. J Geriatr Phys Ther. 2017 Oct/Dec;40(4):223-226. doi: 10.1519/JPT.0000000000000102. PMID: 27805924.

7. Podsiadlo D, Richardson S. The timed “Up & Go”: a test of basic functional mobility for frail elderly persons. J Am Geriatr Soc. 1991 Feb;39(2):142–8. doi: 10.1111/j.1532-5415.1991.tb01616.x. PMID: 1991946.

8. Huang SL, Hsieh CL, Wu RM, Tai CH, Lin CH, Lu WS. Minimal detectable change of the timed “up & go” test and the dynamic gait index in people with Parkinson disease. Phys Ther. 2011 Jan;91(1):114–21. doi: 10.2522/ptj.20090126. Epub 2010 Oct 14. Erratum in: Phys Ther. 2014 Jul;94(7):1056. PMID: 20947672.

9. Giovannoni G, van Schalkwyk J, Fritz VU, Lees AJ. Bradykinesia akinesia inco-ordination test (BRAIN TEST): an objective computerised assessment of upper limb motor function. J Neurol Neurosurg Psychiatry. 1999;67(5):624–9. doi:10.1136/jnnp.67.5.624

10. Noyce AJ, Nagy A, Acharya S, Bestwick JP, Dobbs AJ, Lees AJ, et al. Bradykinesia-akinesia incoordination test: validating an online keyboard test of upper limb function. PLoS One. 2014;9(4):e96260. doi:10.1371/journal.pone.0096260

11. Hasan H, Burrows M, Athauda DS, Roussakis AA, Roussakis AA, Warner TT, et al. The BRadykinesia Akinesia INcoordination (BRAIN) Tap Test: Capturing the Sequence Effect. Mov Disord Clin Pract. 2019;6(6):462–9. doi:10.1002/mdc3.12798

12. Akram N, Li H, Ben-Joseph A, Budu C, Gallagher DA, Bestwick JP, Schrag A, Noyce AJ, Simonet C. Developing and assessing a new web-based tapping test for measuring distal movement in Parkinson’s disease: a Distal Finger Tapping test. Sci Rep. 2022 Jan 10;12(1):386. doi: 10.1038/s41598-021-03563-7. PMID: 35013372; PMCID: PMC8748736.

13. Mak MK, Pang MY. Balance confidence and functional mobility are independently associated with falls in people with Parkinson’s disease. J Neurol. 2009 May;256(5):742–9. doi: 10.1007/s00415-009-5007-8. Epub 2009 Feb 25. PMID: 19240961.

14. Buysse DJ, Reynolds CF 3rd, Monk TH, Berman SR, Kupfer DJ. The Pittsburgh Sleep Quality Index: a new instrument for psychiatric practice and research. Psychiatry Res. 1989 May;28(2):193–213. doi: 10.1016/0165-1781(89)90047-4. PMID: 2748771.

15. Teruel-Hernández E, López-Pina JA, Souto-Camba S, Báez-Suárez A, Medina-Ramírez R, Gómez-Conesa A. Improving Sleep Quality, Daytime Sleepiness, and Cognitive Function in Patients with Dementia by Therapeutic Exercise and NESA Neuromodulation: A Multicenter Clinical Trial. Int J Environ Res Public Health. 2023 Nov 6;20(21):7027. doi: 10.3390/ijerph20217027. PMID: 37947583; PMCID: PMC10650908.

16. Jenkinson C, Fitzpatrick R, Peto V, Greenhall R, Hyman N. The Parkinson’s Disease Questionnaire (PDQ-39): development and validation of a Parkinson’s disease summary index score. Age Ageing. 1997 Sep;26(5):353–7. doi: 10.1093/ageing/26.5.353. PMID: 9351479.

17. Horváth K, Aschermann Z, Kovács M, Makkos A, Harmat M, Janszky J, Komoly S, Karádi K, Kovács N. Changes in Quality of Life in Parkinson’s Disease: How Large Must They Be to Be Relevant? Neuroepidemiology. 2017;48(1-2):1–8. doi: 10.1159/000455863. Epub 2017 Feb 4. PMID: 28161701.

18. Roberts SB, Bonnici DM, Mackinnon AJ, Worcester MC. Psychometric evaluation of the Hospital Anxiety and Depression Scale (HADS) among female cardiac patients. Br J Health Psychol. 2001 Nov;6(Part 4):373–383. doi: 10.1348/135910701169278. PMID: 12614511.

19. Hagell P, Höglund A, Reimer J, Eriksson B, Knutsson I, Widner H, Cella D. Measuring fatigue in Parkinson’s disease: a psychometric study of two brief generic fatigue questionnaires. J Pain Symptom Manage. 2006 Nov;32(5):420–32. doi: 10.1016/j.jpainsymman.2006.05.021. PMID: 17085268.

20. Hurst H, Bolton J. Assessing the clinical significance of change scores recorded on subjective outcome measures. J Manipulative Physiol Ther. 2004 Jan;27(1):26–35. doi: 10.1016/j.jmpt.2003.11.003. PMID: 14739871.

21. Griffiths RI, Kotschet K, Arfon S, Xu ZM, Johnson W, Drago J, et al. Automated assessment of bradykinesia and dyskinesia in Parkinson’s disease. J Parkinsons Dis. 2012;2(1):47–55. doi:10.3233/JPD-2012-11071

22. Kotschet K, Johnson W, McGregor S, Hall C, Gschwandtner U, Horne M. Daytime sleep in Parkinson’s disease measured by episodes of immobility. Parkinsonism Relat Disord. 2014;20(6):578–83. doi:10.1016/j.parkreldis.2014.02.011

23. Horne MK, McGregor S, Bergquist F. An objective fluctuation score for Parkinson’s disease. PLoS One. 2015;10(4):e0124522. doi:10.1371/journal.pone.0124522

24. Braybrook M, O’Connor S, Churchward P, Perera T, Farzanehfar P, Horne M. An Ambulatory Tremor Score for Parkinson’s Disease. J Parkinsons Dis. 2016;6(4):723–31. doi:10.3233/JPD-160898

25. Horne M, Kotschet K, McGregor S. The clinical validation of objective measurement of movement in Parkinson’s disease. CNS. 2016;2(1):16–23.

26. Shokouhi N, Khodakarami H, Fernando C, Osborn S, Horne M. Accuracy of step count estimations in Parkinson’s disease can be predicted using ambulatory monitoring. Front Aging Neurosci. 2022;14:904895. doi:10.3389/fnagi.2022.904895

27. McGregor S, Churchward P, Soja K, Baque E, Macfarlane MD, McInerney J, et al. The use of accelerometry as a tool to measure disturbed nocturnal sleep in Parkinson’s disease. NPJ Parkinsons Dis. 2018;4:1. doi:10.1038/s41531-017-0038-9

28. Klingelhoefer L, Rizos A, Sauerbier A, McGregor S, Martinez-Martin P, Reichmann H, et al. Night-time sleep in Parkinson’s disease – the potential use of Parkinson’s KinetiGraph: a prospective comparative study. Eur J Neurol. 2016 Aug;23(8):1275–88. doi:10.1111/ene.13015

